# The effect of herpes zoster vaccination at different stages of the disease course of dementia: Two quasi-randomized studies

**DOI:** 10.1101/2024.08.23.24312457

**Authors:** Min Xie, Markus Eyting, Christian Bommer, Haroon Ahmed, Pascal Geldsetzer

## Abstract

The varicella zoster virus, a neurotropic herpesvirus, has been hypothesized to play a role in the pathophysiology of dementia, such as through neuroinflammatory processes or intracerebral vasculopathy. Using unique natural experiments, our group has previously found that live-attenuated herpes zoster (HZ) vaccination reduced the incidence of new diagnoses of dementia in both Wales and Australia. To inform further research and ultimately clinical care, it is crucial to understand at which stage of the disease course of dementia the HZ vaccine has its effect. Representing the two opposing ends of the dementia disease course as it can be ascertained from electronic health record data, the aims of this study were twofold: to determine the effect of HZ vaccination on i) new diagnoses of mild cognitive impairment (MCI) among individuals without any record of cognitive impairment, and ii) deaths due to dementia among individuals living with dementia. Our approach took advantage of the fact that at the time of the start date (September 1 2013) of the HZ vaccination program in Wales, individuals who had their eightieth birthday just after this date were eligible for HZ vaccination for one year whereas those who had their eightieth birthday just before were ineligible and remained ineligible for life. This eligibility rule created comparison groups just on either side of the September 2 1933 date-of-birth eligibility threshold who differed in their age by merely a week but had a large difference in their probability of receiving HZ vaccination. The key strength of our study is that these comparison groups should be similar in their health characteristics and behaviors except for a minute difference in age. We used regression discontinuity analysis to estimate the difference in our outcomes between individuals born just on either side of the date-of-birth eligibility threshold for HZ vaccination. Our dataset consisted of detailed country-wide electronic health record data from primary care in Wales, linked to hospital records and death certificates. We restricted our dataset to individuals born between September 1 1925 and September 1 1942. Among our study cohort of 282,557 without any record of cognitive impairment at baseline, HZ vaccination eligibility and receipt reduced the incidence of a new MCI diagnosis by 1.5 (95% CI: 0.5 – 2.9, p=0.006) and 3.1 (95% CI: 1.0 – 6.2, p=0.007) percentage points over nine years, respectively. Similarly, among our study cohort of 14,350 individuals who were living with dementia at baseline, being eligible for and receiving HZ vaccination reduced deaths due to dementia by 8.5 (95% CI: 0.6 – 18.5, p=0.036) and 29.5 (95% CI: 0.6 – 62.9, p=0.046) percentage points over nine years, respectively. Except for dementia, HZ vaccination did not have an effect on any of the ten most common causes of morbidity and mortality among adults aged 70 years and older in Wales in either of our two study cohorts. The protective effects of HZ vaccination for both MCI and deaths due to dementia were larger among women than men. Our findings suggest that the live-attenuated HZ vaccine has benefits for the dementia disease process at both ends of the disease course of dementia.

## Introduction

Given the key role of neuroinflammation in the development and progression of dementia(*1*), it is conceivable that neurotropic viruses could be a factor that causes or accelerates the dementia disease process. Neurotropic herpesviruses have thus far received the greatest research attention in this regard(*2–4*) because they remain latent for life in the nervous system after primary infection, are more likely to reactivate with increasing age, and can cause encephalitis(*5*). Recently, several findings have further spurred interest in neurotropic herpesviruses, including the observation that they can seed β-amyloid in mice(*6*) and that the Epstein Barr Virus appears to be a causative factor in the development of Multiple Sclerosis(*7*).

The neurotropic herpesvirus (the varicella zoster virus) that causes chickenpox and shingles has recently been linked to amyloid deposition and aggregation of tau proteins(*8*), as well as cerebrovascular disease that resembles the patterns commonly seen in Alzheimer’s disease, such as small to large vessel disease, ischemia, infarction, and hemorrhage(*9–14*). Reducing clinical and subclinical reactivations of the virus through herpes zoster (HZ) vaccination might, thus, have a beneficial impact on the development or progression of dementia. Moreover, as has been detailed recently elsewhere(*15*), it is possible that HZ vaccination, and potentially vaccinations in older age more generally, act on the dementia disease process through a pathogen-independent immune mechanism. Such an effect would add to the growing body of evidence suggesting that vaccines frequently have broader health benefits beyond their intended target(*16–18*). Of importance with respect to this present study, these beneficial off-target effects have often been found to be far stronger among female than male individuals(*17*), and for live-attenuated rather than other types of vaccines(*16–19*).

In a recent preprint(*20*), we were able to take advantage of a unique quasi-randomization in Wales to provide evidence on the effect of HZ vaccination on new dementia diagnoses that is more likely to be causal than the previously existing associational evidence(*21–30*). This opportunity arose because the UK National Health Service rolled out the live-attenuated HZ vaccine (Zostavax, Merck) using strict date of birth-based eligibility rules(*31*). These rules resulted in an increase in the probability of ever receiving the HZ vaccine of almost 50 percentage points between individuals who differed in their age by merely a week across the date of birth-based eligibility threshold for the vaccination program. We, thus, had the opportunity to compare dementia incidence between eligible and ineligible groups of individuals who were not expected to differ in their characteristics other than a difference in age of merely a few weeks and a large difference in ever receiving the HZ vaccine. We found that HZ vaccination averted an estimated one in five new dementia diagnoses over a seven-year follow-up period. Crucially, unlike the previously existing associational evidence(*21–29*), this study is not subject to the fundamental concern in associational studies that those who opt to be vaccinated differ from those who do not in a variety of characteristics that are difficult to measure(*32*). Most recently, taking advantage of a similar date of birth-based rollout of HZ vaccination in Australia, we have shown that this protective effect for new diagnoses of dementia from HZ vaccination also exists in the Australian population(*33*).

To guide further research in this area and, ultimately, inform appropriate clinical care, it is critical to understand at which stage of the disease course of dementia the HZ vaccine has its benefit. Our previous analyses in Wales and Australia have left this question unanswered. The aims of this study, therefore, were twofold: to determine the effect of HZ vaccination on i) new diagnoses of mild cognitive impairment (MCI) among individuals without any record of cognitive impairment, and ii) deaths due to dementia among individuals living with dementia. In the absence of more widespread testing for amyloid β and tau pathology during the study period (2013 to 2022), these two aims represent the two opposite ends of the disease course of dementia (considering the limitations in ascertaining different disease stages in electronic health record data). Thus, observing a beneficial effect from HZ vaccination in both aims would suggest that the vaccine appears to act across the entire disease course of dementia.

## Methods

### The herpes zoster vaccine rollout in Wales

Starting on September 1 2013, the National Health Service ([NHS], the United Kingdom’s single-payer single-provider healthcare system(*34*)) in Wales made the live-attenuated HZ vaccine (Zostavax, Merck) available to a catch-up cohort of individuals using a staggered rollout system based on specific date-of-birth eligibility thresholds(*31*). Individuals who did not yet have their 80^th^ birthday on the start date of the program (i.e., born on or after September 2 1933) were eligible for one year. By contrast, those who had their 80^th^ birthday prior to the program start date (i.e., born before September 2 1933) never became eligible. A more detailed description of the rollout is provided in **Supplement Text S1**.

### Data source

This study used the Secure Anonymised Information Linkage (SAIL) Databank(*35*, *36*). This databank provides detailed electronic health record data from primary care in the NHS through the Welsh Longitudinal General Practice dataset(*37*), which contains data on diagnoses, clinical signs and observations, symptoms, laboratory tests and results (via the Welsh Results Report Service(*38*)), procedures performed (including vaccinations), prescribed medications, and administrative items(*39*). Using individuals’ unique NHS number, SAIL links this primary care dataset to a series of databases. For our study, these databases consisted of the Welsh Demographic Service Dataset(*40*), the Patient Episode Database for Wales (containing hospital-based inpatient care data)(*41*), the Outpatient Database for Wales (containing specialist-based ambulatory care data)(*42*), the Welsh Cancer Intelligence and Surveillance Unit (containing data on care for cancer)(*43*), and the Annual District Death Extract (containing cause-of-death data)(*44*). Our data included individuals’ date of birth in weeks (with weeks starting on a Monday). A detailed description of each dataset is provided in **Supplement Text S2**.

### Study cohorts and follow-up period

We restricted our dataset to all individuals born between September 1 1925 and September 1 1942 who were ever registered with a primary care provider in Wales, which is the case for over 98% of Wales’s adult population(*45*), and who were alive and residing in Wales as of the start date of the HZ vaccination program (September 1 2013). Given that each patient in our dataset had a unique NHS number, we were able to follow patients over time even if they changed primary care provider. We defined one study cohort for each of our two aims. For determining the effect of HZ vaccination on the incidence of MCI, we excluded patients whose electronic health record data suggested any cognitive impairment at any time prior to the start date of the HZ vaccination program. To do so, we used the code list for cognitive impairment published by Moran et al. (also shown in **Supplement Materials**)(*46*), which consists of detailed Read codes for any symptoms, signs, and diagnoses relating to cognitive impairment, such as disturbances of memory, orientation, concentration, or reasoning, as well as formal diagnoses of MCI and dementia. For determining the effect of HZ vaccination on the occurrence of deaths due to dementia, we restricted our analysis cohort to those patients with a diagnosis of dementia made at any time prior to the start date of the HZ vaccination program. This cohort is henceforth referred to as patients living with dementia at baseline. The Read and ICD codes used to define dementia (as well as all other diagnoses used in this study) are provided in **Supplement Materials**.

The follow-up period for all primary analyses was nine years, starting on September 1 2013 (the start date of the HZ vaccination program) and ending on August 31 2022. In secondary analyses, we show all results when using follow-up periods from one to nine years in one-year increments.

### Exposure and outcome definition

The exposure was eligibility for HZ vaccination based on one’s date of birth. As shown in **Supplement Figs. S1** and **S2**, most eligible patients (especially in the first two eligibility cohorts of the phased rollout, which are the focus of our analysis) in each of our two study populations took up HZ vaccination during their first year of eligibility.

For determining the effect of HZ vaccination on MCI, our primary outcome was MCI as defined by a record of a Read code (see **Supplement Materials**) for MCI in our electronic health record data. As robustness check, we required that the first diagnosis of MCI not be followed by a new dementia diagnosis within three and within six months to examine the sensitivity of our findings to the possibility of a patient with mild-to-moderate dementia being falsely classified as having MCI. For determining the effect of HZ vaccination on deaths due to dementia, our primary outcome was defined as dementia being named as the underlying (i.e., primary) cause of death in the patient’s death certificate (see **Supplement Materials** for ICD-10 codes used). We defined dementia as dementia of any type because of our reduced statistical power when studying less common outcomes, as well as the neuropathological overlap between dementia types and difficulty in distinguishing dementia types clinically(*47–49*). Dates of deaths were for the date of death registration as opposed to occurrence, whereby the median delay between death occurrence and registration in Wales in the years from 2001 to 2021 was five days(*50*).

We used all-cause mortality among patients living with dementia at baseline as a secondary outcome. The rationale for analyzing this secondary outcome was that if HZ vaccination reduced deaths due to dementia, it will be important to ascertain whether this effect led to an increase in remaining life expectancy (in which case we would also observe a reduction in all-cause mortality) or merely to the replacement of dementia as the underlying cause of death on the death certificate with the mentioning of another cause (in which case we would observe no effect on all-cause mortality). The Read and ICD codes used to define all our outcomes, including those used as baseline balance checks and in negative control outcome analyses, as well as HZ vaccination are provided in **Supplement Materials**.

### Statistical analysis

The two authors who analyzed the data (M.E. and M.X.) conducted all parts of the analysis independently, compared their results, and, in the case of any discrepancies, agreed on the preferred coding approach through discussion.

#### Regression discontinuity analysis

Patients born immediately before versus immediately after September 2 1933 would be expected to be exchangeable (i.e., similar in observable and unobservable characteristics) with each other except for their probability of receiving HZ vaccination (as a result of their eligibility status for HZ vaccination). Our analysis approach was guided by this expectation.

Regression discontinuity (RD) is a well-established method for causal effect estimation for such threshold-based exposure assignments(*51*). This technique estimates the outcome probability for individuals just on either side of the September 2 1993 date-of-birth eligibility threshold. As per recommended practice for RD(*52–54*), we used a mean squared error (MSE)-optimal bandwidth with robust bias-corrected standard errors(*55*), and assigned a higher weight to observations closer to either side of the September 2 1933 date-of-birth eligibility threshold using triangular kernel weights. The MSE-optimal bandwidth was calculated separately for each combination of study cohort and outcome definition. We used local linear regression because it is the recommended and most reliable approach for RD analyses even when the relationship between date of birth and the outcome in the entire dataset is exponential(*56*). However, in robustness checks, we also analyzed our data using local quadratic instead of linear regression. Higher polynomial regressions are not recommended for RD(*56*). In addition, we verified that our results were not dependent on the choice of i) bandwidth (by using bandwidth choices of 0.50, 0.75, 1.25, 1.50, 1.75, and 2.00 times the MSE-optimal bandwidth), and ii) grace period (i.e., the time since the index date after which follow-up time is considered to begin to allow for the time needed for a full immune response to develop after vaccine administration). In an additional robustness check, we adjusted the follow-up period to account for the staggered rollout of the HZ vaccination program. Thus, instead of starting the follow-up period for all individuals on September 1 2013, we started the follow-up period for each individual on the date on which they first became eligible for HZ vaccination (as detailed in **Supplement Text S1**). We added cohort fixed effects in these analyses to control for the one-to two-year (depending on the program year) differences between eligibility cohorts in the start of their follow-up period.

For all outcomes, we estimated both the effect of being eligible for HZ vaccination based on one’s date of birth (the intent-to-treat [ITT] effect), as well as the effect of actually receiving HZ vaccination (the complier average causal effect [CACE]). To estimate the CACE, we followed standard practice for RD by implementing a so-called fuzzy RD(*54*). While still comparing individuals just on either side of the date-of-birth eligibility threshold, fuzzy RD corrects the effect estimates for the fact that a proportion of eligible individuals did not receive the vaccine and a small proportion of ineligible individuals did receive the vaccine. Fuzzy RD is implemented by using an instrumental variable approach(*54*). In our analysis, the instrumental variable was a binary indicator for whether or not an individual was eligible for HZ vaccination (i.e., born on or after versus born before September 2 1933). This analysis, therefore, adjusted the effect size for being eligible for HZ vaccination for the magnitude of the abrupt change in the probability of receiving HZ vaccination at the September 2 1933 threshold. Importantly, fuzzy RD does not compare eligible vaccine recipients with eligible vaccine non-recipients because these groups likely have different health characteristics and behaviors (and, thus, confounding is likely).

Given its implementation using local linear regression, RD yields absolute as opposed to relative effect estimates. All regression equations used in our analyses are shown in **Supplement Text S3**.

#### Testing for confounding from a competing intervention

The key advantage of our quasi-randomization approach is that a confounding variable can only bias our analysis if the variable changes abruptly at precisely the September 2 1933 date-of-birth threshold(*52*, *53*). Thus, confounding bias is unlikely unless another intervention existed that used the identical date-of-birth eligibility threshold (i.e., September 2 1933) as the HZ vaccination program. We investigated whether such a competing intervention was likely to exist in four ways.

First, if another intervention that used the identical date-of-birth eligibility threshold had been implemented prior to the HZ vaccination program, then we may expect to observe differences in patients’ health characteristics or past uptake of preventive health services at the time of the start date of the HZ vaccination program. We, therefore, tested for differences (using the same RD approach as we used for our primary outcomes) across the September 2 1933 date-of-birth threshold in the prevalence of i) diagnoses made at any time prior to September 1 2013 for each of the ten most common causes of disability-adjusted life years (DALYs) and mortality among the age group 70+ years in Wales(*57*), and ii) indicators of past preventive health services uptake. The indicators of past preventive health services uptake available in our data were influenza vaccine receipt in the 12 months preceding program start, receipt of the pneumococcal vaccine as an adult, current statin use (defined as a new or repeat prescription of a statin in the 12 months preceding program start), current use of an antihypertensive medication (defined as a new or repeat prescription of an antihypertensive drug in the 12 months preceding program start), and breast cancer screening participation (defined as the proportion of women with a record of referral to, attendance at, or a report from “breast cancer screening” or mammography at any time prior to the start date of the HZ vaccination program). The codes used to define each of these variables is provided in **Supplement Materials**.

Second, if a dementia-specific intervention that used the identical date-of-birth eligibility threshold had been implemented before the HZ vaccination program, then we may expect to observe differences in the incidence of our outcomes across the September 2 1933 threshold prior to the start date of the HZ vaccination program. We, thus, conducted the identical analysis as for our primary outcomes except for starting the follow-up period nine years prior to the start date (September 1 2013) of the HZ vaccination program.

Third, if an annual intervention used September 2 as a date-of-birth eligibility criterion, then we may expect to observe significant differences in our outcomes at the September 2 date-of-birth threshold for birth years other than 1933. We, thus, implemented the same analyses as we did for the September 2 1933 threshold for the September 2 threshold of each of the three years of birth preceding and succeeding 1933 (i.e., date-of-birth thresholds of September 2 1930, September 2 1931, September 2 1932, September 2 1934, September 2 1935, and September 2 1936). To ensure that our analyses at these additional date-of-birth thresholds compared individuals of the same age range as in our primary analyses, we shifted the start and end date of the follow-up period to the same extent as the date-of-birth threshold. To maintain the same follow-up period in all comparisons, we, therefore, had to use a follow-up period of six as opposed to nine years. As an example, when comparing individuals across the September 2 1930 threshold, we started the follow-up period on September 1 2010 and ended the follow-up period on August 31 2016.

Fourth, unless another intervention that used the identical date-of-birth eligibility threshold was specifically designed to affect MCI and dementia only, we may expect to see an effect of such an intervention on health outcomes other than MCI, dementia, and deaths due to dementia. We, thus, conducted the same analysis as for our primary outcomes but for diagnoses of, and deaths due to, each of the ten leading causes of DALYs and mortality in Wales for the age group 70+ years(*57*), as well as indicators of preventive health services uptake available in our data. The indicators of preventive health services uptake were breast cancer screening among women (defined as a record of referral to, attendance at, or a report from “breast cancer screening” or mammography at any time after the start date of the HZ vaccination program), and, for the 12 months after the start of the HZ vaccination program, uptake of influenza vaccination as well as any prescription of a statin or antihypertensive medication.

#### Testing for ascertainment bias

If healthcare seeking for episodes of shingles constituted an important opportunity for the health system to identify previously undiagnosed MCI, then our analysis for the effect of HZ vaccination on MCI could suffer from ascertainment bias. We conducted three analyses to investigate whether this potential ascertainment bias was likely to represent an important source of bias in this study. First, if shingles episodes were an important opportunity for the health system to detect previously undiagnosed chronic conditions, then we may expect to observe an effect of HZ vaccination not only on MCI but also on other common chronic conditions. As described in the preceding section, we, therefore, implemented the same RD analysis as for MCI but for each of the ten leading causes of DALYs and mortality in Wales in 2019 for the age group 70+ years as outcomes(*57*). Second, if healthcare utilization for shingles had an important bearing on the health system’s ability to diagnose MCI, then we may expect that controlling for indicators of healthcare utilization during the follow-up period would attenuate our effect estimates. We, therefore, adjusted our regressions for the number of primary care visits, outpatient visits, hospital admissions, and influenza vaccinations received during our nine-year follow-up period. Third, patients who frequently visit their primary care provider may be more likely to be (whether formally or informally) screened for MCI. An analysis in this cohort of patients should, therefore, be less susceptible to ascertainment bias. We, thus, also implemented our analysis when restricting our study population to the sample of those 135,712 (48.0% of the analysis cohort for our primary analyses for MCI) patients who had made at least one visit to their primary care provider during each of the five years preceding the start of the HZ vaccination program.

#### Triangulation via a different quasi-experimental approach

We used a second quasi-experimental approach, namely a difference-in-differences (DID) analysis, to further investigate the robustness of our RD findings. After restricting our sample to patients born between March 1 1926 and February 28 1934, we implemented our DID approach by dividing our sample into yearly birth cohorts centered around September 1. We then divided each yearly birth cohort into a pre-September birth “season” and a post-September birth season. The pre-September birth season was, thus, defined as the six-months period of March 1 to August 31 and the post-September birth season as the six-months period from September 1 to February 28 of the succeeding year. Our DID model tested whether the difference in outcomes across birth seasons was different for the 1933/1934 birth cohort compared to other yearly birth cohorts. The rationale for our DID was that HZ vaccination eligibility only differed between the two birth seasons in the 1933/1934-cohort but not in other yearly birth cohorts. The DID approach naturally adjusts for any potential systematic differences between pre-September and post-September birth seasons. The regression equations for this DID approach are detailed in **Supplement Text S3**.

Importantly, our DID did not rely on the continuity assumption (i.e., the assumption that potential confounding variables do not abruptly change at exactly the September 2 1933 date-of-birth eligibility threshold) made by RD. Instead, our DID relied on the assumption that had the HZ vaccination program not existed, then the difference in our outcomes between the pre-and post-September birth seasons would have been the same in the 1933/1934-cohort as in other yearly cohorts. A strength of our approach is that we were able to investigate whether this assumption was likely to be met by testing whether there were significant between-birth-season differences in our outcomes in cohorts other than the 1933/1934-cohort. We did identify such significant differences for MCI among patients without a record of cognitive impairment at baseline, but not for deaths due to dementia among patients living with dementia at baseline. Details are provided in **Supplement Fig. S3**. We, therefore, used the DID approach only when analyzing the effect of HZ vaccination on deaths due to dementia.

#### Effect heterogeneity by gender

In our previous analysis for the effect of HZ vaccination on new diagnoses of dementia(*20*), we found a stronger effect among women than men. We, therefore, tested for an effect heterogeneity by gender in both of our aims. To do so, in addition to analyzing the effect among women and men separately, we implemented an interaction model that estimated the difference in effects by gender (the regression equations for this analysis are provided in **Supplement Text S3**).

### Ethics

Approval was granted by the Information Governance Review Panel (IGRP, application number: 1306), which oversees and approves applications to use the SAIL databank. All analyses were approved and considered minimal risk by the Stanford University Institutional Review Board on June 9 2023 (protocol number: 70277).

## Results

### Sample characteristics

Our dataset consisted of 304,940 individuals born between September 1 1925 and September 1 1942 who were alive and residing in Wales as of September 1 2013. Of these individuals, 282,557 did not have a record of any cognitive impairment prior to September 1 2013 and were, thus, included in our study cohort for analyzing the effect of HZ vaccination on MCI. Our study cohort for analyzing the effect of HZ vaccination on deaths due to dementia consisted of the 14,350 individuals in our dataset who had received a diagnosis of dementia prior to September 1 2013. The sample characteristics of each of these two cohorts are shown in **Table 1**.

**Table 1.**
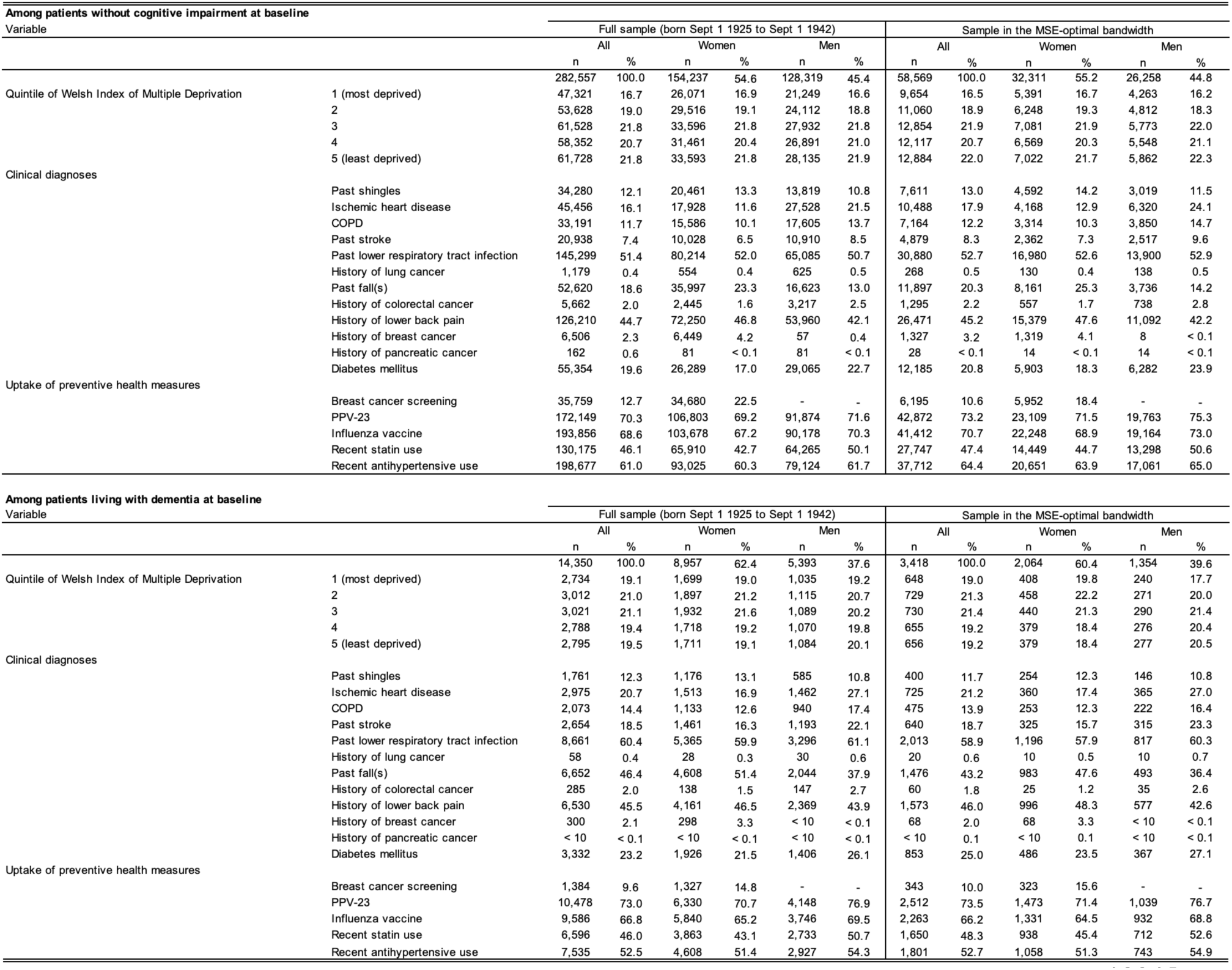
Sample characteristics at baseline of the two study cohorts in our analysis^1,2,3,4,5^. ^1^ The baseline date was September 1 2013 (the start date of the HZ vaccination program). ^2^ The length of the MSE-optimal bandwidth was 95.1 weeks for patients without cognitive impairment at baseline and 97.5 weeks for patients living with dementia at baseline. ^3^ Deciles of the Welsh Index of Multiple Deprivation (WIMD) were calculated based on the 2011 WIMD survey(*58*). ^4^ Breast cancer screening was defined as the proportion of women with a record of referral to, attendance at, or a report from “breast cancer screening” or mammography at any time prior to September 1 2013. “PPV-23” was defined as receipt of the pneumococcal vaccine as an adult at any time prior to September 1 2013. “Influenza vaccine” was defined as influenza vaccine receipt in the 12 months preceding September 1 2013. Recent statin and antihypertensive use was defined as a new or repeat prescription of a statin or antihypertensive drug, respectively, in the 12 months preceding September 1 2013. ^5^ Diabetes mellitus referred to both diabetes mellitus type 1 or type 2. Abbreviations: Sept = September; MSE = mean squared error-optimal; COPD = Chronic Obstructive Pulmonary Disease; PPV-23 = Pneumococcal Polysaccharide Vaccine

### A one-week difference in age led to a large difference in HZ vaccination uptake

In both of our study cohorts, a one-week difference in age across the September 2 1933 date-of-birth eligibility threshold resulted in a large difference in the probability of ever receiving HZ vaccination (**Fig. 1**). Specifically, among individuals without any record of cognitive impairment prior to the start of the HZ vaccination program, being born one week after September 2 1933, and thus being eligible for HZ vaccination, led to an abrupt increase in the probability of ever receiving HZ vaccination from 0.0% to 45.9% (p<0.001). The corresponding abrupt increase among patients living with dementia on the start date of the HZ vaccination program was from 0.0% to 28.7% (p<0.001). Thus, in both of our study cohorts, the eligibility rules of the HZ vaccination program created comparison groups born just on either side of the September 2 1933 date-of-birth threshold who were likely similar to each other except for a minor difference in age and a large difference in the probability of receiving HZ vaccination.

**Fig. 1.**
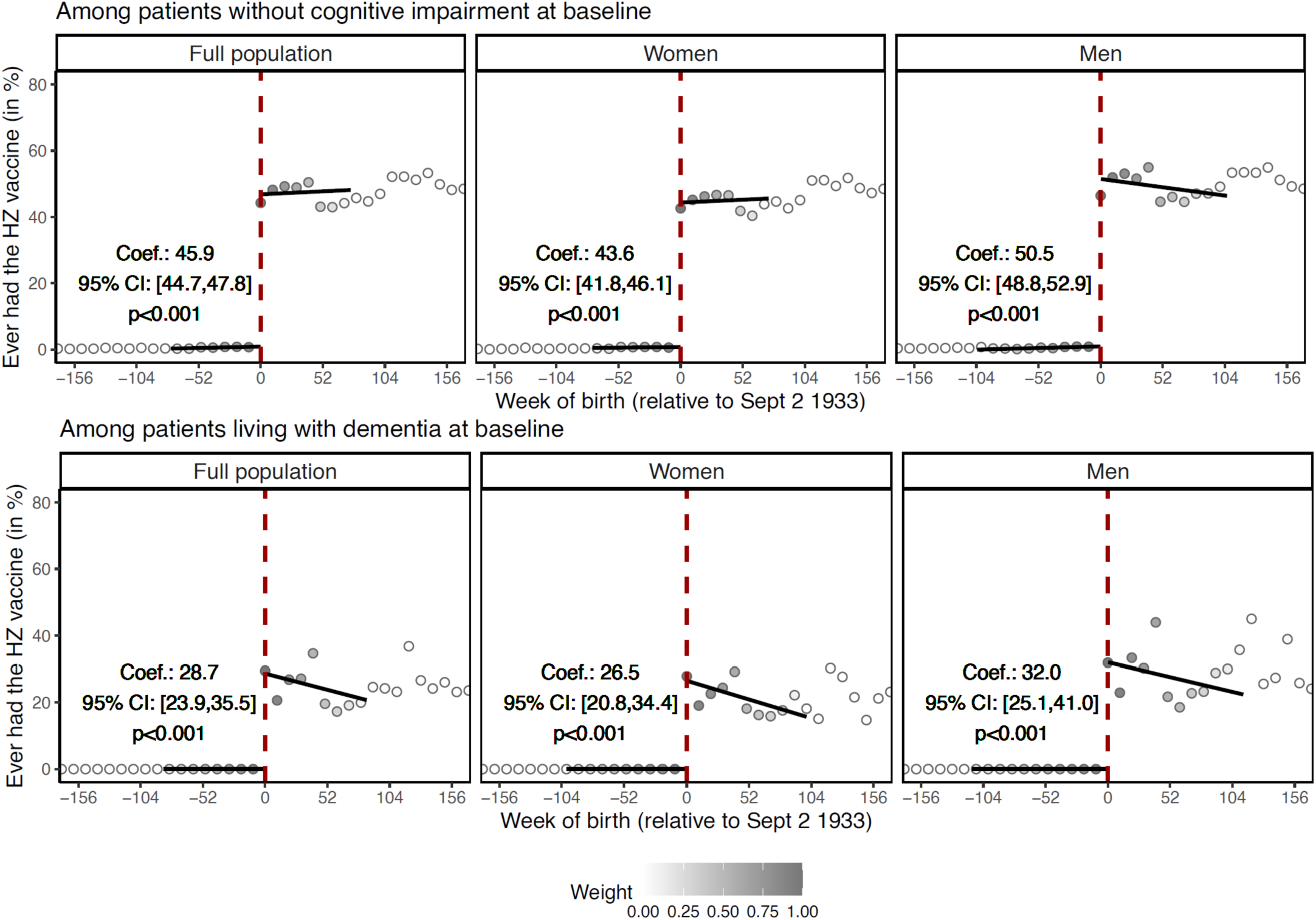
The abrupt change in the probability of receiving HZ vaccination at the September 2 1933 date-of-birth eligibility threshold.^1,2,3,4^. ^1^ “Baseline” refers to the start date of the HZ vaccination program (i.e., September 1 2013). ^2^ Linear regression lines were drawn in the MSE-optimal bandsdwidth only. ^3^ Grey dots show the mean value for each 10-week increment in week of birth. ^4^ The grey shading of the dots is in proportion to the weight that observations from this 10-week increment received in the analysis. Abbreviations: HZ = herpes zoster; Coef. = coefficient; CI = confidence interval; Sept = September

Prior to the start date of the HZ vaccination program, there were no significant differences at the September 2 1933 date-of-birth threshold in the uptake of preventive health services, the prevalence of any of the ten most common causes of DALYs and mortality among adults aged 70+ years in Wales (except ischemic heart disease among those living with dementia at program start), the occurrence of HZ, diagnoses of MCI, and deaths due to dementia (**Supplement Figs. S4** and **S5**).

### The effect of HZ vaccination on new diagnoses of mild cognitive impairment

Among our study cohort of individuals without any record of cognitive impairment prior to the start date of the HZ vaccination program, 20,712 (7.3%) were newly diagnosed with MCI during our nine-year follow-up period. Being born immediately after versus immediately before September 2 1933, and thus being eligible for HZ vaccination, decreased the incidence of new diagnoses of MCI over nine years by 1.5 (95% CI: 0.5 – 2.9, p=0.006) percentage points (**Fig. 2**). Scaled to the proportion of individuals who took up HZ vaccination if they were eligible using the fuzzy RD approach, the effect of actually receiving HZ vaccination was a 3.1 (95% CI: 1.0 – 6.2, p=0.007) percentage point reduction in new diagnoses of MCI over nine years. The effect across different follow-up periods is shown in **Supplement Fig. S6**. Both the effect of being eligible for HZ vaccination and the effect of actually receiving HZ vaccination were robust across different choices of bandwidth, grace period, and functional form (using local quadratic instead of local linear regression), when requiring that a new MCI diagnosis not be followed by a new dementia diagnosis within three and six months, when adjusting for the staggered rollout of the program, when adjusting for indicators of health service utilization, and when restricting the analysis cohort to frequent primary care visitors (**Supplement Fig. S7**).

**Fig. 2.**
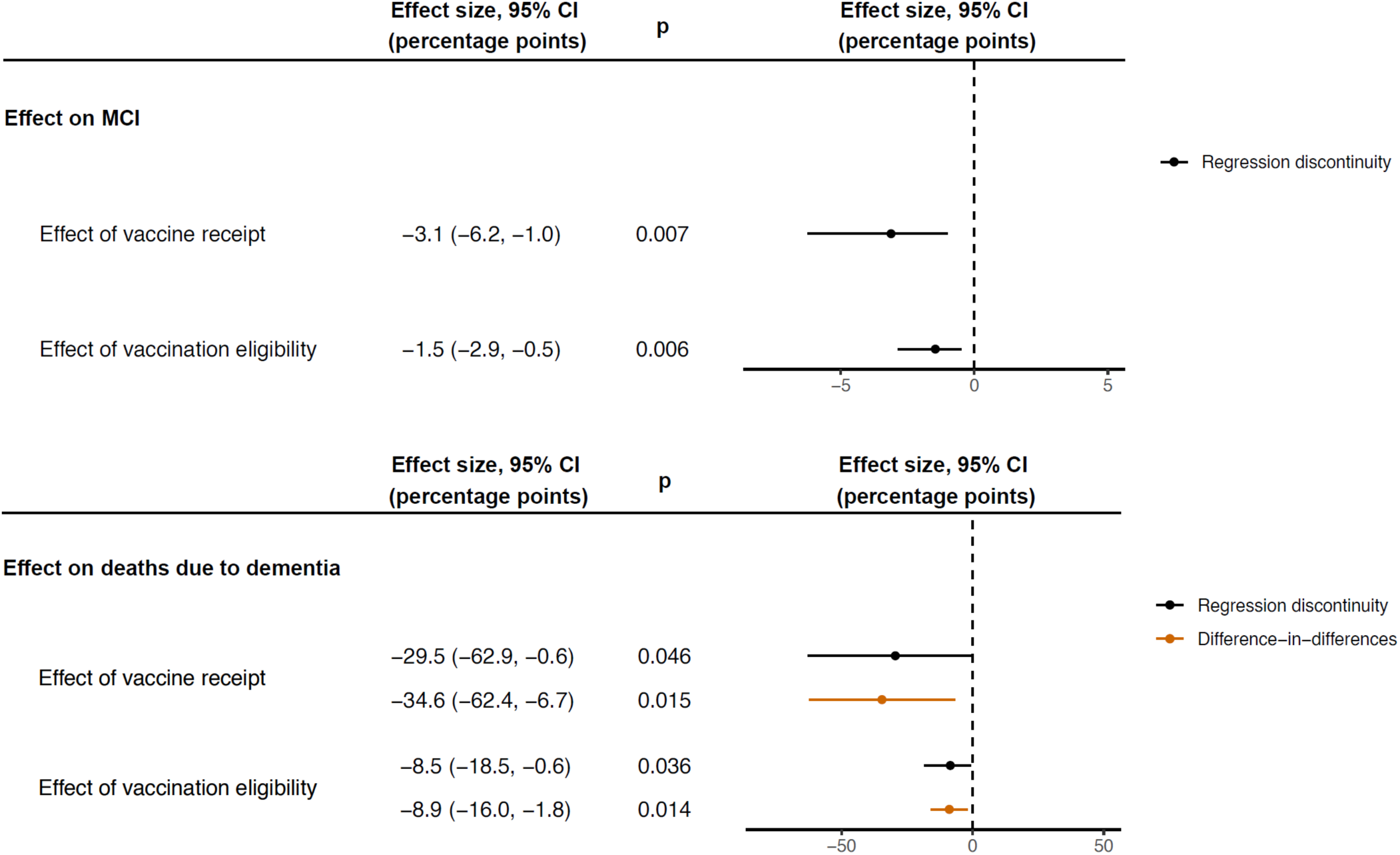
The effect of HZ vaccination on new diagnoses of MCI and deaths due to dementia.^1,2,3^. ^1^ Dots show the point estimate and horizontal bars the 95% confidence interval. ^2^ New diagnoses of MCI were analyzed among a study cohort of patients who did not have any record of cognitive impairment prior to the start date of the HZ vaccination program. ^3^ Deaths due to dementia were analyzed among a study cohort of patients who had received a diagnosis of dementia prior to the start date of the HZ vaccination program. Abbreviations: MCI = mild cognitive impairment; CI = confidence interval

### The effect of HZ vaccination on deaths due to dementia

Among our study cohort of individuals with a diagnosis of dementia received prior to the start date of the HZ vaccination program, 7,049 (49.1%) died due to dementia over the nine-year follow-up period. Being eligible for HZ vaccination (i.e., being born shortly after versus shortly before September 2 1933) decreased the incidence of deaths due to dementia over nine years by 8.5 (95% CI: 0.6 – 18.5, p=0.036) percentage points (**Fig. 2**). The effect of actually receiving HZ vaccination was a 29.5 (95% CI: 0.6 – 62.9, p=0.046) percentage point reduction in deaths due to dementia over nine years. Our DID analysis yielded similar results as our RD analysis (**Fig. 2**). The effect across different follow-up periods is shown in **Supplement Fig. S8**. As for MCI as outcome, the point estimates for the effect of being eligible for HZ vaccination and the effect of actually receiving HZ vaccination were robust across different choices of bandwidth, grace period, and functional form (using local quadratic instead of local linear regression), and when adjusting for the staggered rollout of the program (**Supplement Fig. S9**). In addition, the effect remained significant when adding a dichotomous covariate that indicated whether an individual had been diagnosed with ischemic heart disease prior to the start date of the HZ vaccination program (**Supplement Fig. S9**).

### Testing for confounding

Given the focus of our RD approach on changes in the outcome variable at the date-of-birth eligibility threshold, a confounding variable would only bias our analysis if it changed abruptly at precisely the September 2 1933 date-of-birth threshold. Such bias could arise if another intervention used the identical date-of-birth eligibility threshold (i.e., September 2 1933) as the HZ vaccination program. As detailed in the Methods section, we tested for this possibility in four ways.

First, the September 2 date-of-birth eligibility threshold only had a significant effect on our primary outcomes in the birth year (1933) that was used by the HZ vaccination program, but not in any of the three birth years prior to and after 1933 (**Supplement Fig. S10**). This finding reduces the probability that an annual intervention existed that also used September 2 as a date-of-birth eligibility criterion.

Second, at the time of the start date of the HZ vaccination program, we did not observe systematic differences across the September 2 1933 date-of-birth threshold in past preventive health services uptake nor the prevalence of the ten leading causes of DALYs and mortality among adults aged 70+ years in Wales (**Supplement Fig. S4** and **S5**). Third, there were no differences in the incidence of new MCI diagnoses (in our study cohort for analyzing the effect on MCI) and deaths due to dementia (in our study cohort for analyzing the effect on deaths due to dementia) across the September 2 1933 date-of-birth threshold in the nine years before the start of the HZ vaccination program (**Supplement Fig. S11**). Together, these tests reduce the likelihood that a competing intervention (i.e., another intervention that used the identical date-of-birth eligibility threshold as the HZ vaccination program) existed that was implemented prior to the HZ vaccination program.

Fourth, other than for dementia, we did not observe any significant effects of the September 2 1933 date-of-birth eligibility threshold on diagnoses of each the ten most common causes of DALYs and mortality (in our study cohort for analyzing the effect on MCI) and deaths due to the ten leading causes of mortality (in our study cohort for analyzing the effect on deaths due to dementia) among adults aged 70+ years in Wales over our nine-year follow-up period (**Fig. 3**). Neither did we observe any effects on indicators of preventive health services uptake during our follow-up period (**Fig. 3**). These tests provide evidence against the existence of an intervention that used the identical date-of-birth eligibility threshold as the HZ vaccination program and was not specifically designed to only affect our primary outcomes.

**Fig. 3.**
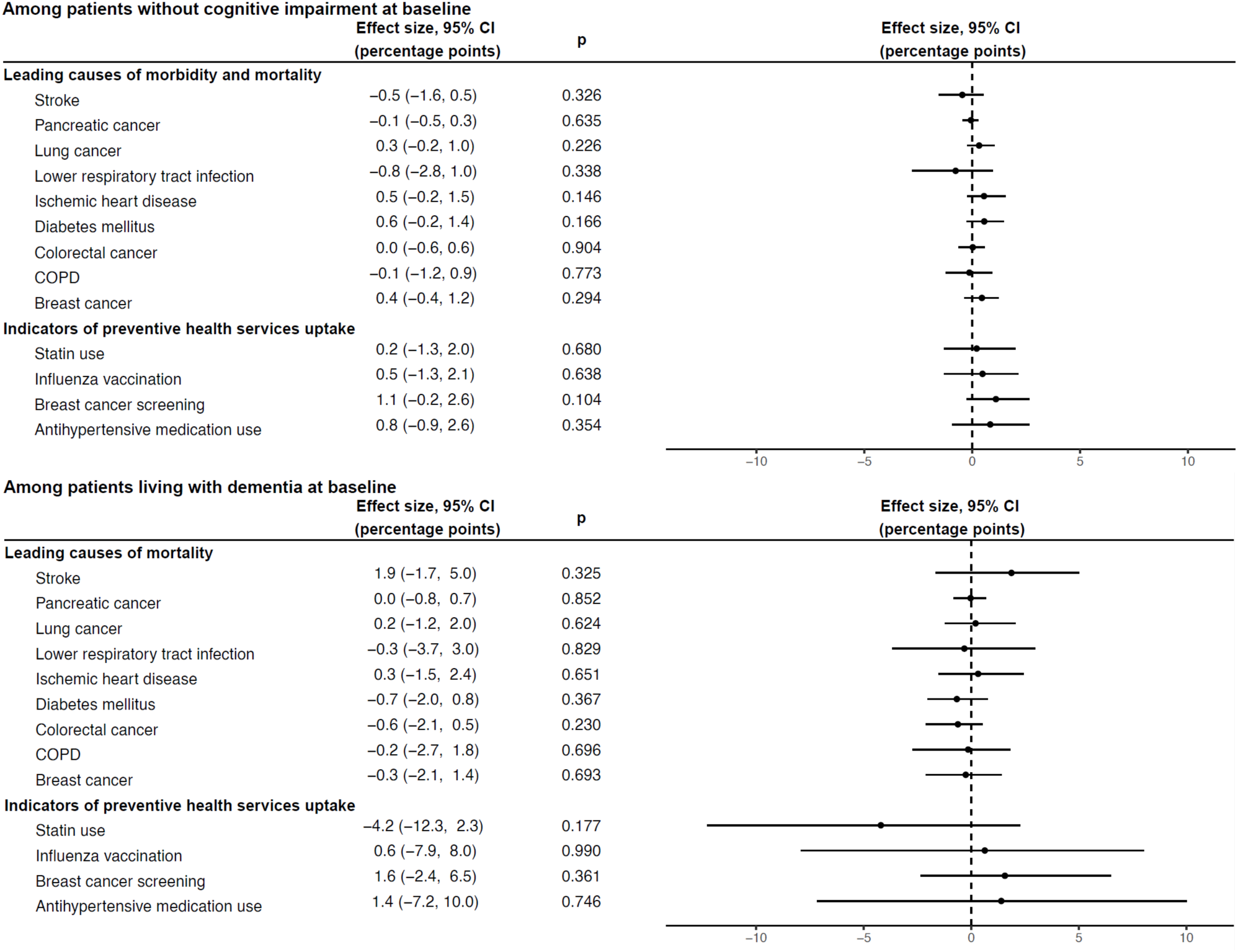
No significant effect of being eligible for HZ vaccination on the leading causes of morbidity and mortality (other than dementia) nor on indicators of preventive health services uptake.^1,2,3,4,5,6,7,8^. ^1^ Baseline refers to the start date (September 1 2013) of the HZ vaccination program. ^2^ Dots show the point estimate and horizontal bars the 95% confidence interval. ^3^ Among patients without a record of cognitive impairment prior to the start date of the HZ vaccination program, the leading causes of morbidity and mortality were the ten (other than dementia) leading causes of DALYs and mortality among adults aged 70+ years in Wales as estimated by the Global Burden of Disease Project(*57*). ^4^ Among patients with a diagnosis of dementia made prior to the start date of the HZ vaccination program, the leading causes of mortality were the ten (other than dementia) leading causes of mortality among adults aged 70+ years in Wales as estimated by the Global Burden of Disease Project(*57*). ^5^ Influenza vaccination was defined as receipt of influenza vaccination at any time in the 12 months after the start date of the HZ vaccination program. ^6^ Statin and antihypertensive medication use was defined as any prescription of these medications during the 12 months after the start date of the HZ vaccination program. ^7^ Breast cancer screening was analyzed among women only. It was defined as a record of referral to, attendance at, or a report from “breast cancer screening” or mammography at any time after the start date of the HZ vaccination program. ^8^ The Read and ICD codes used to define each variable shown in this figure are provided in **Supplement Materials**. Abbreviations: COPD = Chronic Obstructive Pulmonary Disease; CI = Confidence Interval.

### Effect heterogeneity by gender

The effect of HZ vaccination both on reducing new diagnoses of MCI and deaths due to dementia was larger among women than men (**Fig. 4**). Among women, the effects of being eligible for, and actually receiving, HZ vaccination on new diagnoses of MCI were a reduction of 2.2 (95% CI: 0.9 – 4.2, p=0.002) and 4.7 (95% CI: 2.2 – 9.4, p=0.002) percentage points, respectively. The corresponding estimates for the effects on deaths due to dementia among women living with dementia at baseline were a decrease of 13.9 (95% CI: 3.4 – 26.3, p=0.011) and 52.3 (95% CI: 9.2 – 97.9, p=0.018) percentage points. For both MCI and deaths due to dementia, the estimates among men were statistically indistinguishable from zero. Formal interaction tests by gender showed that the interaction was significant for both new MCI diagnoses (p=0.029) and deaths due to dementia (p=0.039) (**Supplement Table S1**). The significant effects among women were robust across different choices of bandwidth, grace period, and functional form (using local quadratic instead of local linear regression), as well as when requiring that a new MCI diagnosis not be followed by a new dementia diagnosis within three and six months, adjusting for the staggered rollout of the program, adjusting for indicators of health service utilization, and restricting the analysis cohort to frequent primary care visitors (**Supplement Fig. S12** and **S13**).

**Fig. 4.**
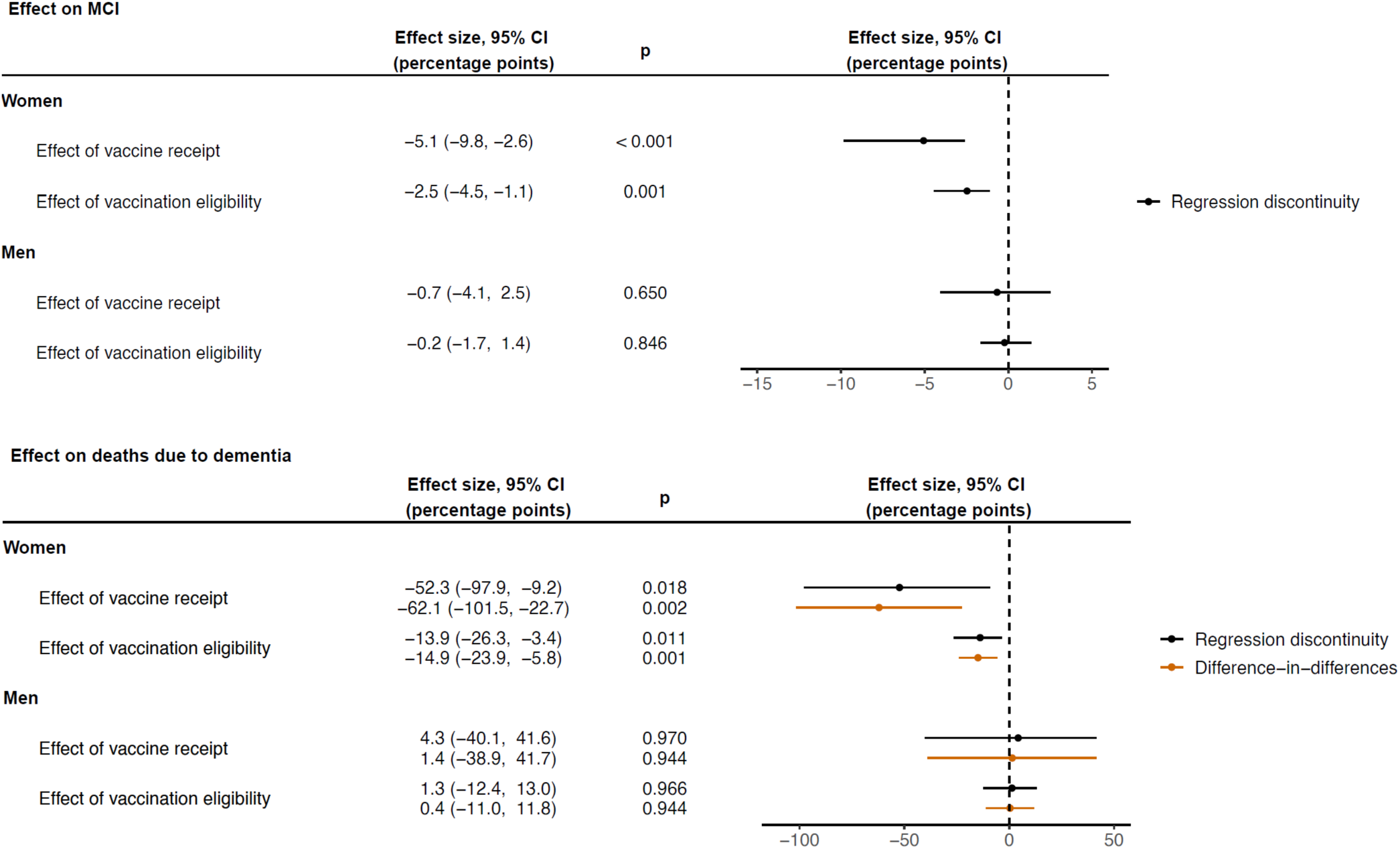
The effect of HZ vaccination on new diagnoses of MCI and deaths due to dementia, separately by gender.^1,2,3^. ^1^ Dots show the point estimate and horizontal bars the 95% confidence interval. ^2^ New diagnoses of MCI were analyzed among a study cohort of patients who did not have any record of cognitive impairment prior to the start date of the HZ vaccination program. ^3^ Deaths due to dementia were analyzed among a study cohort of patients who had received a diagnosis of dementia prior to the start date of the HZ vaccination program. Abbreviations: MCI = mild cognitive impairment; CI = confidence interval

### The effect of HZ vaccination on all-cause mortality

Among patients living with dementia at baseline, being eligible for HZ vaccination (based on being born immediately after versus immediately before September 2 1933) decreased all-cause mortality over our nine-year follow-up period by 6.5 (95% CI: 2.1 – 12.5, p=0.006) percentage points (**Fig. 5**). Adjusted for the proportion who took up HZ vaccination if they were eligible using the fuzzy RD approach, the effect of actually receiving HZ vaccination was a 22.7 (95% CI: 6.5 – 42.8, p=0.008) percentage point reduction in all-cause mortality over nine years. There was no significant effect of HZ vaccination eligibility or receipt on non-dementia deaths. As for deaths due to dementia, the effects of HZ vaccination eligibility and receipt on all-cause mortality were larger among women and statistically indistinguishable from zero among men (**Supplement Fig. S14**). All results were similar for our DID as for our RD analysis. Both among the whole sample and among women only, the effect on all-cause mortality was robust across different choices of bandwidth and grace period, as well as when using local quadratic instead of local linear regression, adjusting for the staggered rollout of the program, and when adding a dichotomous covariate that indicated whether an individual had been diagnosed with ischemic heart disease prior to the start date of the HZ vaccination program (**Supplement Fig. S15** and **S16**).

**Fig. 5.**
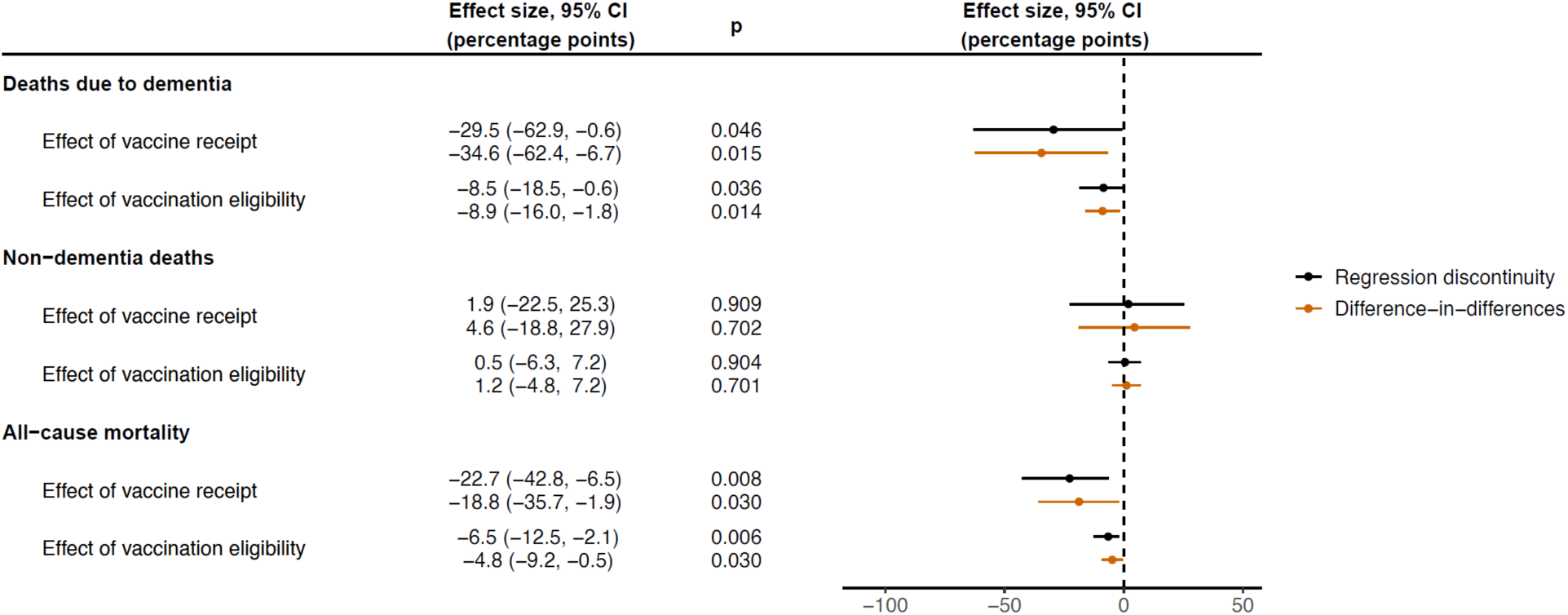
The effect of HZ vaccination on deaths due to dementia, non-dementia deaths, and all-cause mortality.^1,2,3^. ^1^ Dots show the point estimate and horizontal bars the 95% confidence interval. ^2^ Deaths due to dementia were analyzed among a study cohort of patients who had received a diagnosis of dementia prior to the start date of the HZ vaccination program. ^3^ Non-dementia deaths were defined as deaths for which dementia was recorded as neither the underlying nor a contributing cause of death in the death certificate. Abbreviations: CI = confidence interval

## Discussion

We found that HZ vaccination reduced both new diagnoses of MCI among those without any record of cognitive impairment and deaths due to dementia among patients living with dementia. The HZ vaccine, thus, appears to have a beneficial effect at both ends of the disease course of dementia. Whilst our point estimates indicate large effect sizes, the exact magnitude of the effect was difficult to ascertain in this analysis given the wide confidence intervals around our estimates. Lower statistical power compared to standard associational analyses was a result of our focus on individuals born in close proximity to the September 2 1933 date-of-birth threshold.

Our findings suggest that HZ vaccination could be an effective intervention both to prevent or delay MCI and dementia, as well as to reduce disease progression among those already living with dementia. Using the same quasi-randomization approach as in this study, our group has previously shown that HZ vaccination reduced new diagnoses of dementia in both Wales as well as Australia(*20*, *33*). This effect could have been observed as a result of HZ vaccination decreasing the transition from cognitively unimpaired to MCI, from MCI to dementia, or, because of the delay in diagnosing dementia in the health system(*59–63*), from undiagnosed to diagnosed dementia. This present study suggests that the HZ vaccine acts along the entire disease course of dementia and, thus, that our previously reported effects were due to a decrease in the transition along each of these dementia disease stages.

Given that it is a readily available, relatively inexpensive(*64–66*), one-off intervention, the finding that HZ vaccination has a beneficial effect on the dementia disease process would be of great significance to population health, clinical medicine, and dementia research. Thus, confirming our previously reported findings from Wales and Australia is critical. In our view, it is key that such confirmatory studies utilize quasi-randomization opportunities because more standard associational analyses may lead to false confirmations as a result of confounding, such as the healthy vaccinee bias (i.e., the common observation that healthier, more health-motivated, individuals opt to be vaccinated(*32*)). Our quasi-randomized approach is far less vulnerable to these biases because health status, health-related motivation, and other dementia-related characteristics are unlikely to differ between individuals born just before versus just after a specific date-of-birth eligibility threshold. Therefore, in addition to its principal finding that the HZ vaccine appears to act along the entire disease course of dementia, an important contribution of this present study is that it confirms our previously reported findings in two different study populations (those without any record of cognitive impairment and those living with dementia) and using two different dementia-related outcomes (MCI and deaths due to dementia). Deaths with dementia as their primary cause among individuals living with dementia is a particularly opportune outcome in this regard because it is directly related to dementia, but less reliant on a timely diagnosis of dementia in the health system given that dementia is likely to be readily apparent by the time that it is the primary cause of death. Similarly, our secondary outcome of all-cause mortality among individuals living with dementia at baseline is entirely independent of the health system’s process for diagnosing dementia. These mortality outcomes are, thus, largely measured differently than new diagnoses of dementia. In our view, being able to confirm the benefits of HZ vaccination for dementia using a set of outcomes that are all related to dementia, but measured differently, strengthens the evidence that HZ vaccination has an effect on the dementia disease process itself rather than only its measurement (e.g., a diagnostic pathway).

We found that, among individuals living with dementia at baseline, HZ vaccination did not only lead to a decrease in deaths due to dementia but also a reduction in overall mortality. Specifically, we observed a decrease, which was larger among women than men, in both deaths due to dementia and all-cause mortality, but no effect on deaths for which dementia was not mentioned as the underlying or a contributing cause on the death certificate. Our findings, thus, imply that HZ vaccination among individuals living with dementia increased remaining life expectancy. This reduction in deaths due to dementia is unlikely to be a result of averted shingles episodes given that shingles has a low mortality rate(*67*). Instead, this study suggests that the HZ vaccine may slow dementia disease progression. Nonetheless, identifying the exact mechanism for this effect is in our view an important area of future research.

For each of our outcomes, we found that the protective effect of HZ vaccination was larger among women than men. We observed the same gender effect heterogeneity in our previous study in Wales for new diagnoses of dementia(*20*). However, although none of our results were statistically significant among men, we cannot exclude the possibility of substantial beneficial effects among men as well given the width of our confidence intervals. A strong gender effect heterogeneity, with beneficial effects usually being more pronounced among females, has frequently been observed for off-target effects of vaccines(*17*). Our observed effect heterogeneity between women and men may, thus, reflect immunological sex differences.

These immunological sex differences may be pathogen-independent, but could also be specific to the interaction of the immune system with the varicella zoster virus(*68*). The occurrence of shingles has, for instance, been reported to be more common among women in several studies(*69*, *70*). Additionally, it is equally possible that our observed gender effect heterogeneity reflects differences in the pathophysiology of some types of dementia between women and men; an area that has received increasing research interest in recent years(*71–73*).

The key strength of this study is its quasi-randomized design. Prior to our analysis in Wales, all epidemiological studies on the relationship between vaccines and dementia had simply compared vaccine recipients with non-recipients whilst attempting to control for the myriad of characteristics that differ between those who opt to be vaccinated versus those who do not(*21*–*29*). Electronic health record data do not have detailed information on health behaviors, such as physical activity and diet(*74*, *75*), that are known to be linked to other health behaviors (including vaccination) as well as dementia. These studies are, thus, vulnerable to confounding(*76*). Our approach is fundamentally different in that we compare individuals who were ineligible or eligible for HZ vaccination because they were born just before or just after the date-of-birth eligibility threshold (September 2 1933) for HZ vaccination. On average, individuals in Wales born in one week versus merely a week later would not be expected to differ in their health characteristics and behaviors. In the case of the September 2 1933 threshold, however, there was a large difference in the probability of ever receiving the HZ vaccine between these groups who differed in age by merely a week. The eligibility rules of the HZ vaccination program in Wales, thus, created two comparison groups just on either side of the September 2 1933 threshold who were likely similar to each other except for their probability of receiving the intervention of interest (HZ vaccination). As a result, the findings from our analysis are more likely to reflect a causal relationship than those from the associational epidemiological evidence(*21–30*).

The critical advantage of our quasi-randomization approach is that a confounding variable can only bias our analysis if it changes abruptly at precisely the September 2 1933 date-of-birth threshold(*52*, *53*). Such an abrupt change in a confounding variable at the September 2 1933 date-of-birth threshold might exist if there was another intervention that used the identical date-of-birth threshold as its eligibility criterion as the HZ vaccination program. We conducted a series of tests to investigate whether the existence of such an intervention is likely. Specifically, individuals just on either side of the September 2 1933 date-of-birth threshold were well-balanced at baseline in their past preventive health services uptake, prevalence of common causes of morbidity and mortality, and past incidence of MCI and deaths due to dementia.

Similarly, there was no effect of the September 2 1933 date-of-birth threshold on common health outcomes (other than dementia) during our nine-year follow-up period, nor on preventive health services uptake indicators (other than HZ vaccination). Lastly, the September 2 date-of-birth threshold only had an effect on MCI and deaths due to dementia in the birth year (1933) that was used by the HZ vaccination program as its eligibility criterion. Thus, none of our tests suggested the existence of another intervention that also used the date of birth of September 2 1933 as its eligibility criterion. It is in our view also unlikely that HZ vaccination led to increased uptake of other preventive health services (e.g., the uptake of other vaccines at the same visit) because we did not observe any effect of HZ vaccination eligibility on available indicators of preventive health services among older adults in our data.

We believe that our repeated findings from quasi-randomization studies of a beneficial effect of HZ vaccination for the dementia disease process call for further investments into this area of research, not only to confirm the effects but also to elucidate the mechanisms. Several potential mechanisms have already been identified. For instance, there is evidence that reactivations of the varicella zoster virus can lead to long-lasting cognitive impairment through vasculopathy(*77*, *78*), amyloid deposition and aggregation of tau proteins(*8*), neuroinflammation(*11–14*), as well as a pattern of cerebrovascular disease that is similar to that seen in Alzheimer’s disease, such as small to large vessel disease, ischemia, infarction, and hemorrhage(*9–14*). In addition, reactivations of the varicella zoster virus may lead to reactivations of the herpes simplex virus in the brain(*79*). This finding in turn would link HZ to the more extensive literature on the herpes simplex virus as a causative factor in the development of dementia(*3*, *80*). Our repeated finding of a strong effect heterogeneity by gender, however, may also instead point to a pathogen-independent immunomodulatory mechanism(*17*). Some of these potential immune mechanisms have recently been described elsewhere(*15*).

This study has several important limitations. First, given its implementation using local linear regression, RD is only able to reliably estimate absolute as opposed to relative effects(*53*, *54*). Second, we were limited to information contained in electronic health record and death certificate data to define our study cohorts and outcomes. It is likely that there is considerable underascertainment in our data for cognitive impairment (to define the cognitively unimpaired study cohort for our first aim), dementia (to define the study cohort of individuals living with dementia for our second aim), MCI, and deaths due to dementia. Crucially, however, the degree of underascertainment (as well as any delay in making these diagnoses or in changes in data quality over time) is unlikely to differ between individuals born just before versus just after September 2 1933. As such, other than potential underestimation of the true benefits of HZ vaccination on an absolute scale, underascertainment of these variables was unlikely to bias our analysis. Third, we were also limited to the information available in electronic health record data to define the two stages of the disease course of dementia analyzed in this study. We, for instance, had no information on amyloid β or tau pathology or results from detailed neuropsychological assessments. Fourth, the COVID-19 pandemic started within our follow-up period and may have delayed new diagnoses of MCI as well as changed mortality rates among individuals living with dementia. However, as for our first limitation, the pandemic affected individuals just on either side of the September 2 1933 date-of-birth threshold equally and is, thus, unlikely to have introduced bias into our analysis. Fifth, some diagnoses of MCI in our data may have mistakenly been made for individuals who already had mild-to-moderate dementia.

We believe that this is unlikely to be a major limitation of our analysis given that our effect estimates remained significant when requiring that a new diagnosis of MCI not be followed by a new diagnosis of dementia within three and within six months. Sixth, our findings pertain to those age groups born near to the September 2 1933 date-of-birth threshold (primarily those aged 79 to 80 years). We are unable to comment on the effects in other age groups. Seventh, we observed an imbalance in past ischemic heart disease diagnoses across the September 2 1933 threshold among the study cohort for our second aim. We, however, show in the Supplement that our results were robust to any adjustment for this imbalance. In our view, this imbalance was likely a chance finding given that it was the only difference with a p-value less than 0.05 in 36 (18 among each of our two study cohorts) baseline balance tests that we conducted. Lastly, our results pertain to the live-attenuated HZ vaccine (Zostavax, Merck) only, because the newer recombinant HZ vaccine (Shingrix, GSK) was introduced into the NHS after our follow-up period ended(*81*). A recent study of data from the United States that focused on comparing the association of HZ vaccination with dementia when only the older live-attenuated vaccine was available with the association when the newer recombinant vaccine was available, found that the newer vaccine was more strongly associated with a reduced risk of dementia(*30*). The study, however, had to assume that (after matching on select variables available in the electronic health record data) individuals who chose to be vaccinated for shingles when the older, one-dose, vaccine was available were not different in any of their dementia-related characteristics to those who chose to be vaccinated when the far more efficacious(*68*), two-dose, vaccine was available. One limitation, for instance, is therefore the possibility that some individuals may have delayed HZ vaccination to receive the more efficacious vaccine.

In conclusion, this study suggests that HZ vaccination slows or prevents disease progression across the entire disease course of dementia. By taking advantage of the fact that the UK’s National Health Service assigned individuals who differed in their age by just a few weeks to being eligible or ineligible for HZ vaccination based on their date of birth, we were able to generate evidence that is more likely to be causal than those from more standard epidemiological analyses. Our finding that HZ vaccination had a beneficial effect on two different dementia-related outcomes in two different patient samples, and at two opposing ends of the disease course of dementia, thus, provides promising evidence that HZ vaccination may prevent or slow the dementia disease process in a substantial proportion of individuals.

## Data Availability

The data that support the findings of this study are available from the SAIL Databank. Researchers must request access to the data directly from SAIL. The authors have no permission to share the data. All Read and ICD codes to define variables are available in Supplement Materials. All statistical analysis code (in R) will be made available in a publicly accessible GitHub repository upon acceptance of the manuscript for publication.

## Acknowledgements

This study makes use of anonymized data held in the SAIL Databank. The authors are grateful for the continuous advice and support from the SAIL Databank analytical services team throughout all stages of the project. We would also like to acknowledge all the data providers who made anonymized data available for research. The responsibility for the interpretation of the data supplied by SAIL is the authors’ alone. SAIL bears no responsibility for the further analysis or interpretation of their data, over and above that published by SAIL.

## Funding

National Institutes of Health/National Institute on Aging, R01AG084535 (PG)

National Institutes of Health/National Institute of Allergy and Infectious Diseases, DP2AI171011 (PG)

Chan Zuckerberg Biohub investigator award (PG)

## Authors’ contributions

M.X. and M.E. contributed equally to this work. M.X. and M.E. co-devised the methodology, analyzed and processed the data, created data visualizations, interpreted the results, and reviewed and edited the original draft. C.B. co-devised the methodology, interpreted the results, and reviewed and edited the original draft. H.A. interpreted the results, and reviewed and edited the original draft. P.G. conceived the overall project, acquired funding, conceived the study, devised the methodology, was responsible for administration and supervision, interpreted the results, wrote the original draft, and reviewed and edited the original draft.

## Data and materials availability

The data that support the findings of this study are available from the SAIL Databank(*33*). Researchers must request access to the data directly from SAIL. The authors have no permission to share the data. All Read and ICD codes to define variables are available in Supplement Materials. All statistical analysis code (in R) will be made available in a publicly accessible GitHub repository upon acceptance of the manuscript for publication.

