## Supplement for "The effect of herpes zoster vaccination at different stages of the disease course of dementia: Two quasi-randomized studies"

**The file includes:**

Text S1-S3
Figures S1 to S16
Table S1
References

**Text S1: Description of the herpes zoster vaccination rollout in Wales**

The live-attenuated herpes zoster (HZ) vaccine (Zostavax, Merck) was offered to eligible individuals in Wales through primary care providers in the National Health Service (NHS) beginning on September 1 2013. The rollout to eligible individuals was phased. Specifically, individuals were categorized into three cohorts based on their date of birth:

1. A routine cohort consisting of individuals born on or after September 2 1942 who became eligible when they were 70 years old as of September 1 of a given year of the HZ vaccination program;
2. A catch-up cohort consisting of individuals born between September 2 1933 and September 1 1942 who became eligible when they were 79 years (or 78 years, depending on the year of the program) as of September 1 of a given year of the HZ vaccination program and remained eligible until they were 80 years old (again, as of September 1 of a given year of the HZ vaccination program);
3. Individuals born on or before September 1 1933 (i.e., aged 80 years and older as of September 1 2013) who never became eligible.

Our analysis was restricted to adults born between September 1 1925 (88 years old on the start date of the HZ vaccination program) and September 1 1942 (71 years old on the start date of the HZ vaccination program). Those born between September 1 1925 and September 1 1933 were never eligible for HZ vaccination, whereas those born between September 2 1933 and September 1 1942 became eligible as part of the catch-up cohort mentioned above. Specifically, the vaccine was offered to those born:

- Between September 2 1933 and September 1 1934 in the first year of the program (September 1 2013 to August 31 2014);
- Between September 2 1934 and September 1 1936 in the second year (September 1 2014 to August 31 2015);
- Between September 2 1936 and September 1 1937 in the third year (September 1 2015 to August 31 2016);
- Between September 2 1937 and September 1 1938 in the fourth year (September 1 2016 to August 31 2017).

As of April 1 2017, individuals became eligible for the vaccine on their 78$^{th}$ birthday and remained eligible until their 80$^{th}$ birthday.

**Text S2: Description of each dataset used in this study**

Healthcare in Wales is delivered by NHS Wales, which operates within the United Kingdom's single-payer, single-provider healthcare framework(*1*). The Secure Anonymised Information Linkage (SAIL) Databank used in this study was created as part of a collaboration between NHS Wales, the Welsh government, and Swansea University(*2*, *3*). The SAIL Databank contains comprehensive electronic health records from NHS Wales along with a host of social care and administrative datasets, which are all linked at the individual level(*2*, *3*). The following table summarizes the characteristics of the datasets used in this study.

| **Dataset** | **Description** |
|---|---|
| Welsh Demographic Service Dataset (WDSD)(*4*) | This is a registry of all individuals registered with a primary care provider in Wales (over 98% of adults residing in Wales are registered with a primary care provider(*5*)). The following variables from this dataset were used in this study: date of birth in weeks, sex, individuals' anonymized address history (to define residence status in Wales), and the Welsh Index of Multiple Deprivation (the government's measure of relative deprivation for small areas in Wales(*6*)). |
| Welsh Longitudinal General Practice dataset (WLGP)(*7*) | This dataset contains comprehensive electronic health record data from primary care for approximately 80% of primary care practices in Wales and 83% of the Welsh population. All health events are encoded with Read codes(*8*) and include diagnoses, clinical signs and observations, symptoms, medication prescriptions, laboratory tests and results, procedures performed (including vaccinations), and administrative items. This dataset formed the core of our analysis. All variables listed in Supplement Materials that include Read codes in their definition were at least partially ascertained using this dataset. These variables include those needed to ascertain our study cohorts, primary and secondary outcomes, HZ vaccination, diagnoses for our baseline balance checks and negative control outcome analyses, uptake of preventive health services other than HZ vaccination, and healthcare service utilization indicators. |
| Patient Episode Database for Wales (PEDW)(*9*) | This dataset contains electronic health record data for all inpatient and day case activity undertaken in hospitals of NHS Wales as well as data on Welsh residents treated in hospitals that are part of NHS England. Procedures are encoded using OPCS-4 codes(*10*) and diagnoses using ICD-10 codes(*11*). From this dataset, we used dates of admission (to identify the timing of a specific diagnosis and ascertain our healthcare service utilization indicators) and diagnosis codes (to identify a condition first diagnosed in the hospital setting) in this study. |
| Outpatient Database for Wales (OPDW)(*12*) | This dataset contains attendance information for all hospital-based outpatient appointments. We used dates of attendances from this dataset to ascertain our healthcare service utilization indicators. Patients in the NHS generally have to obtain a referral from a primary care provider to access outpatient specialist care(*1*). Thus, |

| | |
|---|---|
| | referrals to, and diagnoses made in, specialist care are contained in the Welsh Longitudinal General Practice dataset. |
| Welsh Cancer Intelligence and Surveillance Unit (WCISU)(*13*) | This is the national cancer registry for Wales, which records all cancer diagnoses given to Welsh residents regardless of the setting in which they were diagnosed or treated. We used dates of diagnoses (to identify the timing of a specific cancer diagnosis) and diagnosis codes (to identify a specific cancer) from this dataset. |
| Annual District Death Extract (ADDE)(*14*) | This is the national registry of all deaths of Welsh residents, including deaths of Welsh residents that occurred outside of Wales. Cause-of-death data from death certificates used ICD-9 coding until 2001 and ICD-10 coding thereafter. Dates for deaths were those on which the death was registered, as opposed to when it occurred. The median delay between death occurrence and registration in England and Wales in the years from 2001 to 2021 was five days(*15*). We used date of death (to identify the timing of death) as well as cause of death (both underlying and contributing causes) in this study. More detailed information on mortality statistics in the United Kingdom is available elsewhere(*16*). |

61

**Text S3: Regression equations**

*Regression discontinuity:*
We used the following model to estimate the effect of being *eligible* for HZ vaccination on our outcomes:

$$(1)\ Y_i = \alpha + \beta_1 D_i + \beta_2 \cdot (WOB_i - c_o) + \beta_3 D_i \cdot (WOB_i - c_0) + \epsilon_i,$$

where $Y_i$ was a dichotomous variable indicating if an individual had the outcome, and $D_i$ was a dichotomous variable indicating eligibility for HZ vaccination (i.e., $D_i$ was equal to one if an individual was born on or after September 2 1933). The term $(WOB_i - c_0)$ and the interaction term $D_i \cdot (WOB_i - c_0)$ adjusted for how many weeks away from the September 2 1933 date-of-birth eligibility threshold an individual was born. These terms fitted two regression lines on either side of the September 2 1933 date-of-birth eligibility threshold whereby the slope of these regression lines could differ on either side of the threshold. The coefficient $\beta_1$ identified the effect of being eligible for HZ vaccination on the outcome.

We used the following model to estimate the effect of *receiving* HZ vaccination on our outcomes:

$$(2)\ Y_i = \theta + \gamma_1 \hat{V}_i + \gamma_2 \cdot (WOB_i - c_o) + \gamma_3 D_i \cdot (WOB_i - c_0) + \varepsilon_i,$$

where $V_i$ was a dichotomous variable indicating if an individual received HZ vaccination and was instrumented by $D_i$. $\hat{V}_i$ was the predicted probability of receiving HZ vaccination based on the estimation of equation (1) with $V_i$ as the left-hand-side variable. $\gamma_1$ was the effect of receiving HZ vaccination. $\theta$ was the constant and $\varepsilon_i$ an error term. All other variables were the same as in regression equation (1). Equation (2), thus, implemented a so-called fuzzy regression discontinuity (RD) design(*17*), which adjusted the effect size for being eligible for HZ vaccination by the size of the abrupt change in the probability of receiving HZ vaccination at the September 2 1933 date-of-birth eligibility threshold. As is common practice for fuzzy RD(*17*), our fuzzy RD achieved this adjustment by using a dichotomous variable that indicated whether or not an individual was eligible to receive HZ vaccination as an instrumental variable for actual receipt of HZ vaccination.

*Difference-in-differences:*
As described in the Methods section in the main manuscript, we implemented our DID approach by dividing our sample into yearly birth cohorts centered around September 1, and then dividing each yearly birth cohort into a pre-September birth "season" (i.e., the six-months period of March 1 to August 31) and a post-September birth season (i.e., the six-months period from September 1 to February 28 of the succeeding year). Our DID approach relied on the assumption that in the absence of the date of birth-based eligibility rule for HZ vaccination, the between-birth-season difference in the outcome would have been the same in the 1933/1934-cohort as in the other yearly birth cohorts. To investigate the validity of this assumption, we estimated the between-birth-season difference in the outcome for each yearly birth cohort, whereby the difference in the 1932/33-cohort served as the reference group. Specifically, we used the following model:

$$(3)\ Y_i = \theta + \sum_{h \neq 1932} (\gamma^h S_i \cdot {C_i}^h) + \eta_m + \eta_c + \epsilon_i,$$

where $Y_i$ was the outcome of patient *i*, and $S_i$ a dichotomous variable indicating that patient *i* was born in the post-September birth season. ${C_i}^h$ indicated whether patient *i* belonged to birth cohort h, with h = 1926/27, 1927/28, …, 1931/32, and 1933/34. $\gamma^h$ identified the between-birth-season difference of the yearly birth cohort h relative to the 1932/33-cohort. $\theta$ was the constant term, $\eta_m$ and $\eta_c$ indicated the birth month and birth cohort fixed-effect, respectively. $\epsilon_i$ was the error term. **Supplement Fig. S3** plots the estimates of $\gamma^h$ with 95% confidence intervals by yearly birth cohort.

We used a two-stage least squares procedure to implement our DID approach. The first stage estimated the change in the probability of receiving HZ vaccination due to the date of birth-based eligibility rule. We used the following model:

$$(4)\ V_i = \theta + \gamma S_i \cdot C_i + \eta_m + \eta_c + \epsilon_i,$$

where $V_i$ was a dichotomous variable indicating whether patient *i* had received HZ vaccination, $S_i$ a dichotomous variable indicating that patient *i* was born in the post-September birth season, and $C_i$ a dichotomous variable indicating that patient *i* was in the 1933/1934 birth cohort. $\gamma$ identified the change in the probability of receiving HZ vaccination as a result of the date of birth-based eligibility rule. $\theta$ was a constant term and $\epsilon_i$ was the error term. $\eta_m$ and $\eta_c$ were the birth month and cohort fixed effect, respectively.

In the second stage, we estimated the effect of receipt of HZ vaccination on our outcomes. We used the following model:

$$(5)\ Y_i = \alpha + \beta \hat{V}_i + \eta_m + \eta_c + \varepsilon_i,$$

where $Y_i$ was the outcome of patient *i*, and $\hat{V}_i$ was the probability of receiving HZ vaccination as a result of the date of birth-based eligibility rule as estimated from the first-stage regression (4). The coefficient $\beta$ identified the effect of HZ vaccination receipt on our outcome. $\theta,\ \eta_m$, and $\eta_c$ were the constant term, birth month, and yearly birth cohort (1926/1927, 1927/1928, …, 1933/1934) fixed effect, respectively. $\varepsilon_i$ was the error term.

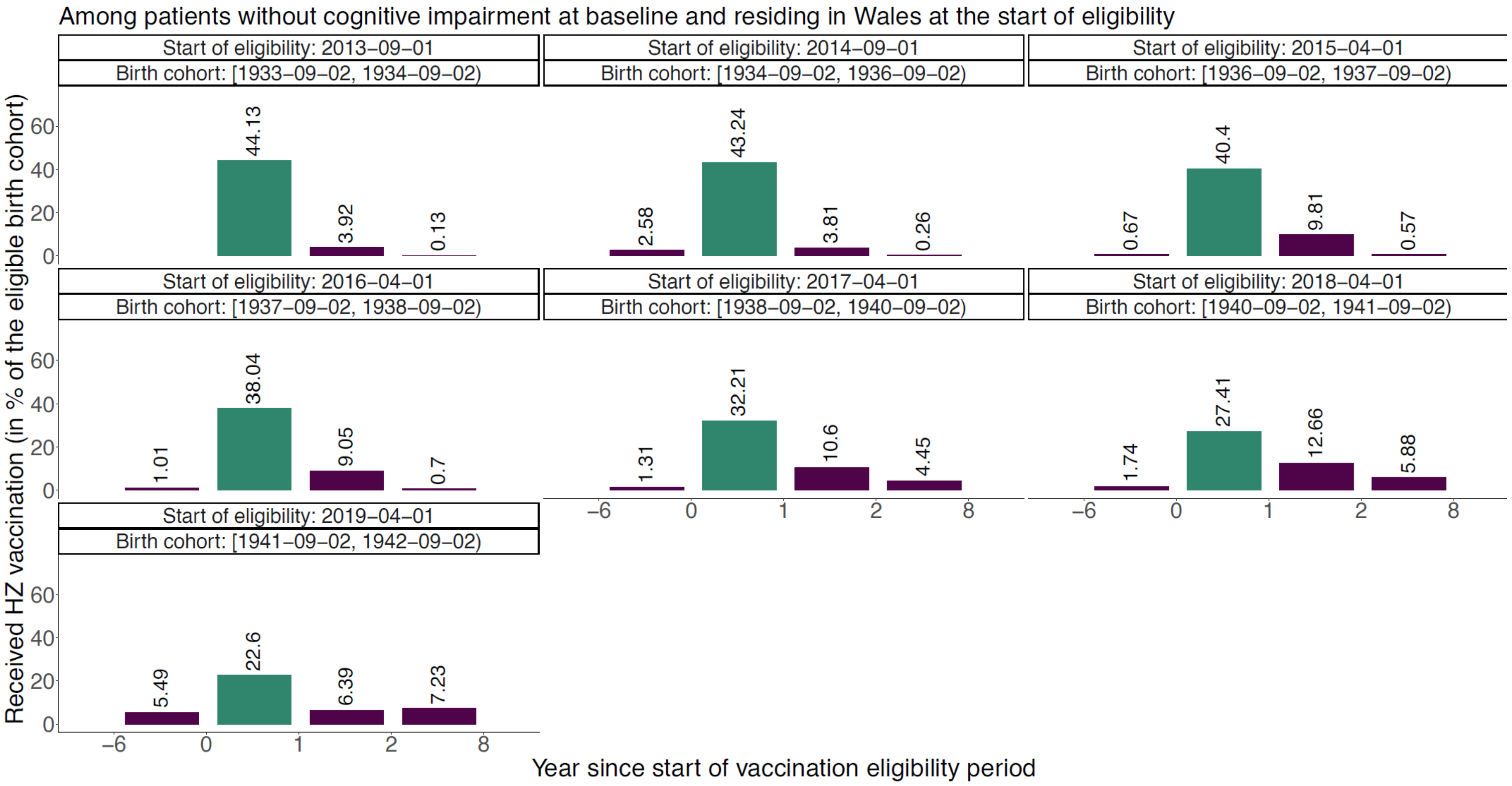


**Figure S1. Uptake of herpes zoster vaccination among patients without cognitive impairment at baseline by birth cohort and year of eligibility.**[1,2,3]

[1] "Baseline" refers to the start date of the HZ vaccination program (i.e., September 1 2013).
[2] The green bar corresponds to the respective first year of eligibility for a particular cohort.
[3] Eligibility years 2 to 8 had to be aggregated to comply with SAIL data publication standards.
Abbreviations: HZ = herpes zoster

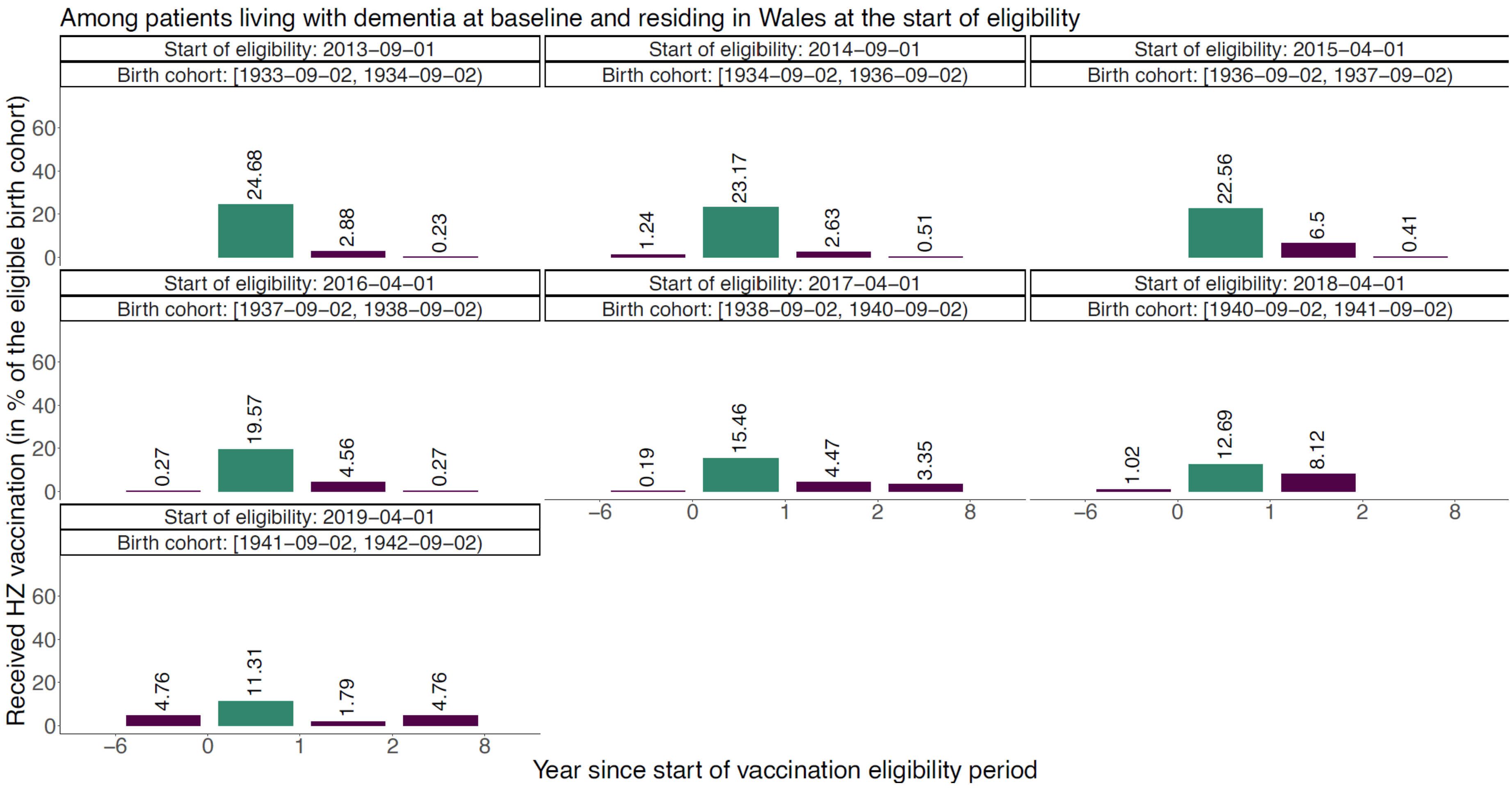


**Figure S2. Uptake of herpes zoster vaccination among patients living with dementia at baseline by birth cohort and year of eligibility.[1,2,3]**

[1] "Baseline" refers to the start date of the HZ vaccination program (i.e., September 1 2013).

[2] The green bar corresponds to the respective first year of eligibility for a particular cohort.

[3] Eligibility years 2 to 8 had to be aggregated to comply with SAIL data publication standards.

Abbreviations: HZ = herpes zoster

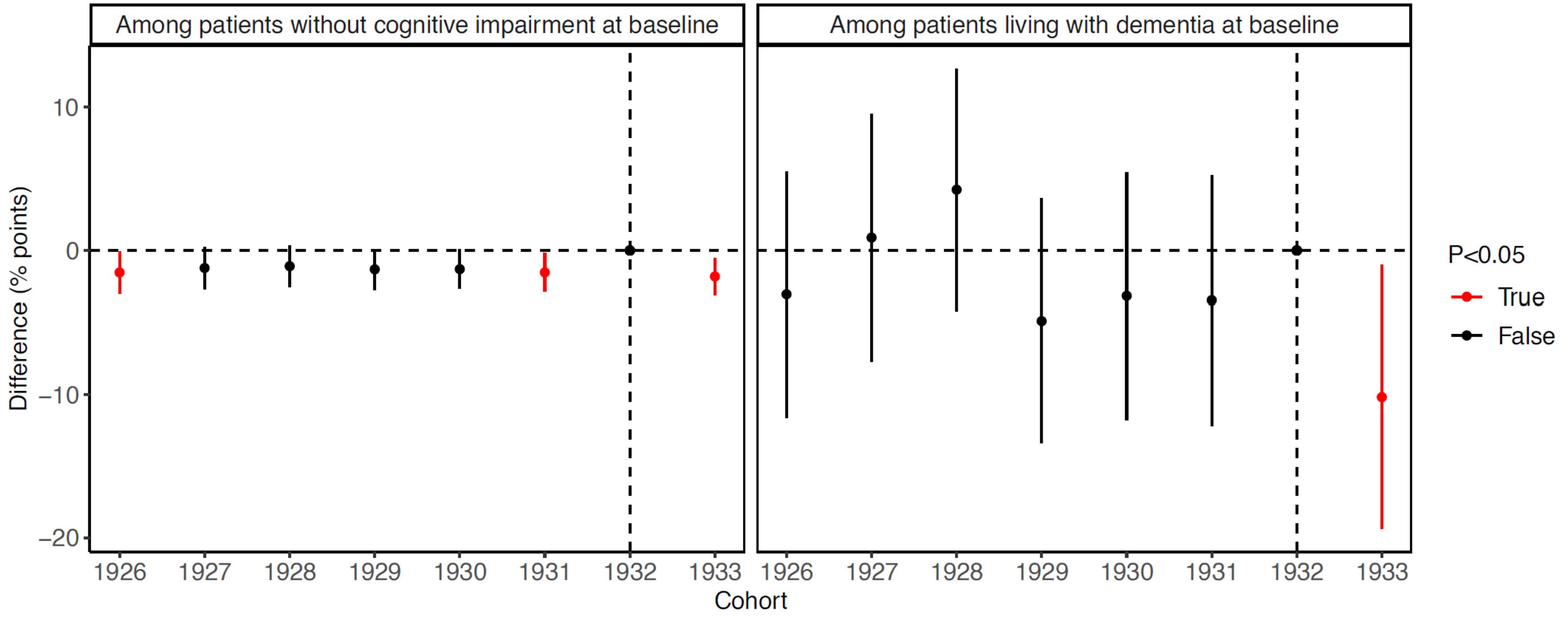


**Figure S3. Difference in the incidence of our primary outcomes between the pre- and post-September birth “seasons” for each yearly birth cohort.**[1,2,3,4,5,6]

[1] “Baseline” refers to the start date of the HZ vaccination program (i.e., September 1 2013).

[2] “Difference” refers to the difference between the pre- and post-September birth “seasons”. The pre-September birth season was defined as the six-months period of March 1 to August 31 of a given year and the post-September birth season as the six-months period from September 1 to February 28 of the succeeding year.

[3] The outcome was new diagnoses of mild cognitive impairment for the study cohort among patients without cognitive impairment at baseline, and deaths due to dementia for the study cohort among patients living with dementia at baseline.

[4] The 1932/1933-cohort served as the reference cohort.

[5] Statistically significant differences (indicated in red) for birth-year cohorts other than the 1933/1934-cohort indicate that the assumption needed for an unbiased difference-in-differences analysis may not be met.

[6] The regression equations for this analysis are detailed in Text S3.

**Among patients without cognitive impairment at baseline**

| | Effect size, 95% CI (percentage points) | p |
|---|---|---|
| **Leading causes of morbidity and mortality** | | |
| Stroke | –0.2 (–1.2, 0.8) | 0.645 |
| Shingles | 0.1 (–1.2, 1.4) | 0.887 |
| Pancreatic cancer | 0.0 (–0.1, 0.1) | 0.926 |
| Mild cognitive impairment | –0.2 (–1.1, 0.4) | 0.390 |
| Lung cancer | 0.1 (–0.2, 0.3) | 0.493 |
| Lower respiratory tract infection | 0.2 (–1.7, 1.9) | 0.909 |
| Lower back pain | –0.9 (–3.0, 0.9) | 0.299 |
| Ischemic heart disease | –0.7 (–2.2, 0.4) | 0.173 |
| Falls | 0.4 (–0.9, 2.0) | 0.462 |
| Diabetes mellitus | 0.0 (–1.5, 1.5) | 0.999 |
| Dementia | 0.5 (–0.3, 1.4) | 0.174 |
| Colorectal cancer | 0.4 (–0.1, 1.2) | 0.113 |
| COPD | 0.0 (–1.1, 1.3) | 0.933 |
| Breast cancer | 0.2 (–0.7, 1.0) | 0.766 |
| **Indicators of preventive health services uptake** | | |
| Statin use | 0.6 (–0.9, 2.6) | 0.351 |
| PPV vaccination | –0.1 (–2.0, 1.3) | 0.696 |
| Influenza vaccination | 0.0 (–1.5, 1.4) | 0.918 |
| Breast cancer screening | –0.5 (–2.5, 1.5) | 0.626 |
| Antihypertensive medication use | 1.2 (–0.7, 3.0) | 0.208 |

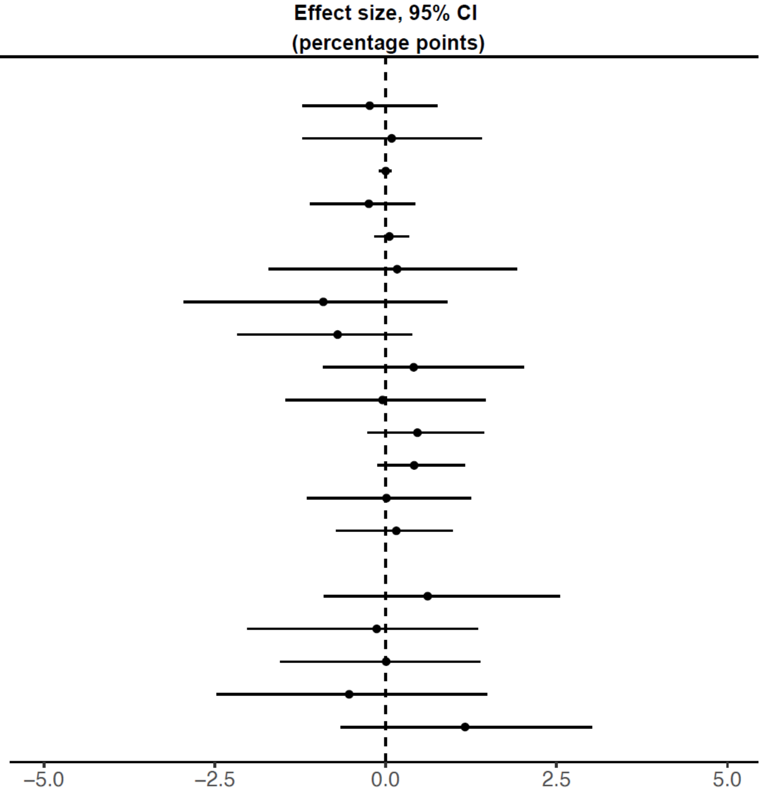


**Figure S4. Baseline balance checks among patients without cognitive impairment at baseline, using diagnoses of the ten most common causes of disability-adjusted life years and mortality among adults aged 70+ years in Wales as well as indicators of preventive health services uptake.**[1,2,3,4,5,6]

[1] "Baseline" refers to the start date of the HZ vaccination program (i.e., September 1 2013).

[2] We used diagnoses of the ten most common causes of disability-adjusted life years (DALYs) and mortality among the age group 70+ years in Wales as estimated by the 2019 Global Burden of Disease study(*18*).

[3] Statin use was defined as a new or repeat prescription of a statin in the 12 months preceding September 1 2013. PPV vaccination was defined as receipt of the PPV vaccine as an adult at any time prior to September 1 2013. Influenza vaccination was defined as receipt of the influenza vaccine in the 12 months preceding September 1 2013. Breast cancer screening was defined as the proportion of women with a record of referral to, attendance at, or a report from "breast cancer screening" or mammography at any time prior to September 1 2013. Antihypertensive medication use was defined as a new or repeat prescription of an antihypertensive drug in the 12 months preceding September 1 2013.

[4] The study cohorts for the baseline balance checks for mild cognitive impairment and dementia did not exclude individuals with any record of cognitive impairment prior to September 1 2013. For all other baseline balance checks, the study cohort was defined as detailed in the Methods section.

[5] Dots show the point estimate and horizontal bars the 95% confidence interval.
[6] The codes used to define each condition are listed in Supplement Materials.
Abbreviations: CI = confidence interval; COPD = Chronic Obstructive Pulmonary Disease; PPV = Pneumococcal Polysaccharide Vaccine

**Among patients living with dementia at baseline**

| | Effect size, 95% CI (percentage points) | p |
|---|---|---|
| **Leading causes of morbidity and mortality** | | |
| Stroke | 0.8 (−4.8, 7.3) | 0.691 |
| Pancreatic cancer | 0.2 (−0.1, 0.5) | 0.173 |
| Lung cancer | 0.5 (−0.2, 1.3) | 0.130 |
| Lower respiratory tract infection | −4.3 (−12.1, 1.8) | 0.148 |
| Lower back pain | 1.2 (−6.8, 10.5) | 0.680 |
| Ischemic heart disease | −8.5 (−15.8, −2.4) | 0.008 |
| Falls | −3.1 (−10.4, 4.4) | 0.424 |
| Diabetes mellitus | −0.6 (−7.2, 6.8) | 0.960 |
| Deaths due to dementia | −5.2 (−17.7, 6.7) | 0.375 |
| Colorectal cancer | −0.3 (−2.2, 1.8) | 0.859 |
| COPD | 0.4 (−4.5, 5.1) | 0.888 |
| Breast cancer | 2.3 (−0.7, 6.3) | 0.121 |
| **Indicators of preventive health services uptake** | | |
| Statin use | −1.9 (−9.8, 5.2) | 0.547 |
| PPV vaccination | −1.2 (−7.3, 4.5) | 0.633 |
| Influenza vaccination | −1.3 (−6.1, 3.0) | 0.502 |
| Breast cancer screening | 6.5 (−0.8, 16.8) | 0.076 |
| Antihypertensive medication use | 3.2 (−5.2, 11.8) | 0.447 |

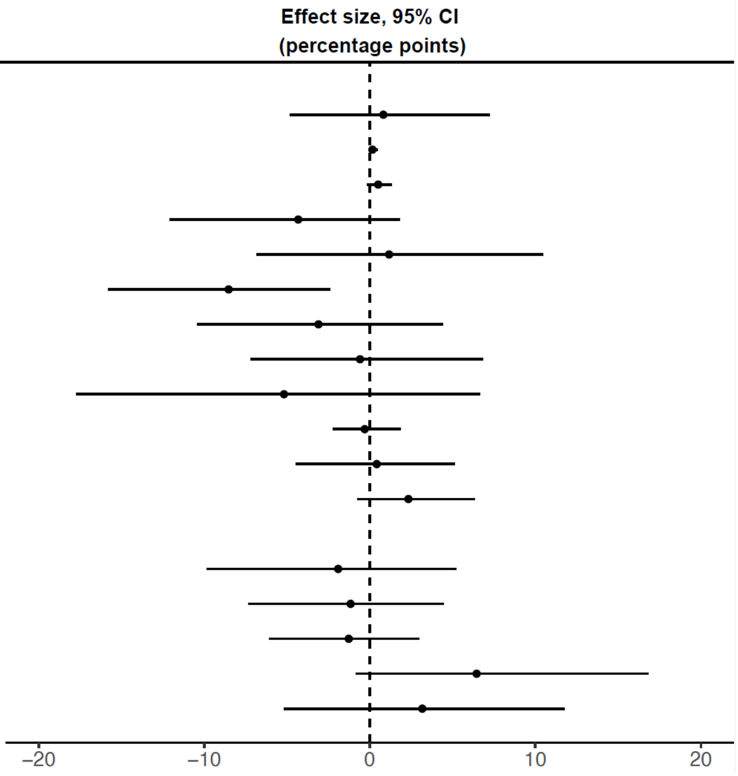


**Figure S5. Baseline balance checks among patients living with dementia at baseline, using diagnoses of the ten most common causes of disability-adjusted life years and mortality among adults aged 70+ years in Wales as well as indicators of preventive health services uptake.**[1,2,3,4,5,6]

[1] "Baseline" refers to the start date of the HZ vaccination program (i.e., September 1 2013).

[2] We used diagnoses of the ten most common causes of disability-adjusted life years (DALYs) and mortality among the age group 70+ years in Wales as estimated by the 2019 Global Burden of Disease study(*18*).

[3] Statin use was defined as a new or repeat prescription of a statin in the 12 months preceding September 1 2013. PPV vaccination was defined as receipt of the PPV vaccine as an adult at any time prior to September 1 2013. Influenza vaccination was defined as receipt of the influenza vaccine in the 12 months preceding September 1 2013. Breast cancer screening was defined as the proportion of women with a record of referral to, attendance at, or a report from "breast cancer screening" or mammography at any time prior to September 1 2013. Antihypertensive medication use was defined as a new or repeat prescription of an antihypertensive drug in the 12 months preceding September 1 2013.

[4] The study cohort for the baseline balance check for deaths due to dementia was not restricted to individuals who were alive on September 1 2013. For all other baseline balance checks, the study cohort was defined as detailed in the Methods section.

[5] Dots show the point estimate and horizontal bars the 95% confidence interval.

[6] The codes used to define each condition are listed in Supplement Materials.
Abbreviations: CI = confidence interval; COPD = Chronic Obstructive Pulmonary Disease; PPV = Pneumococcal Polysaccharide Vaccine

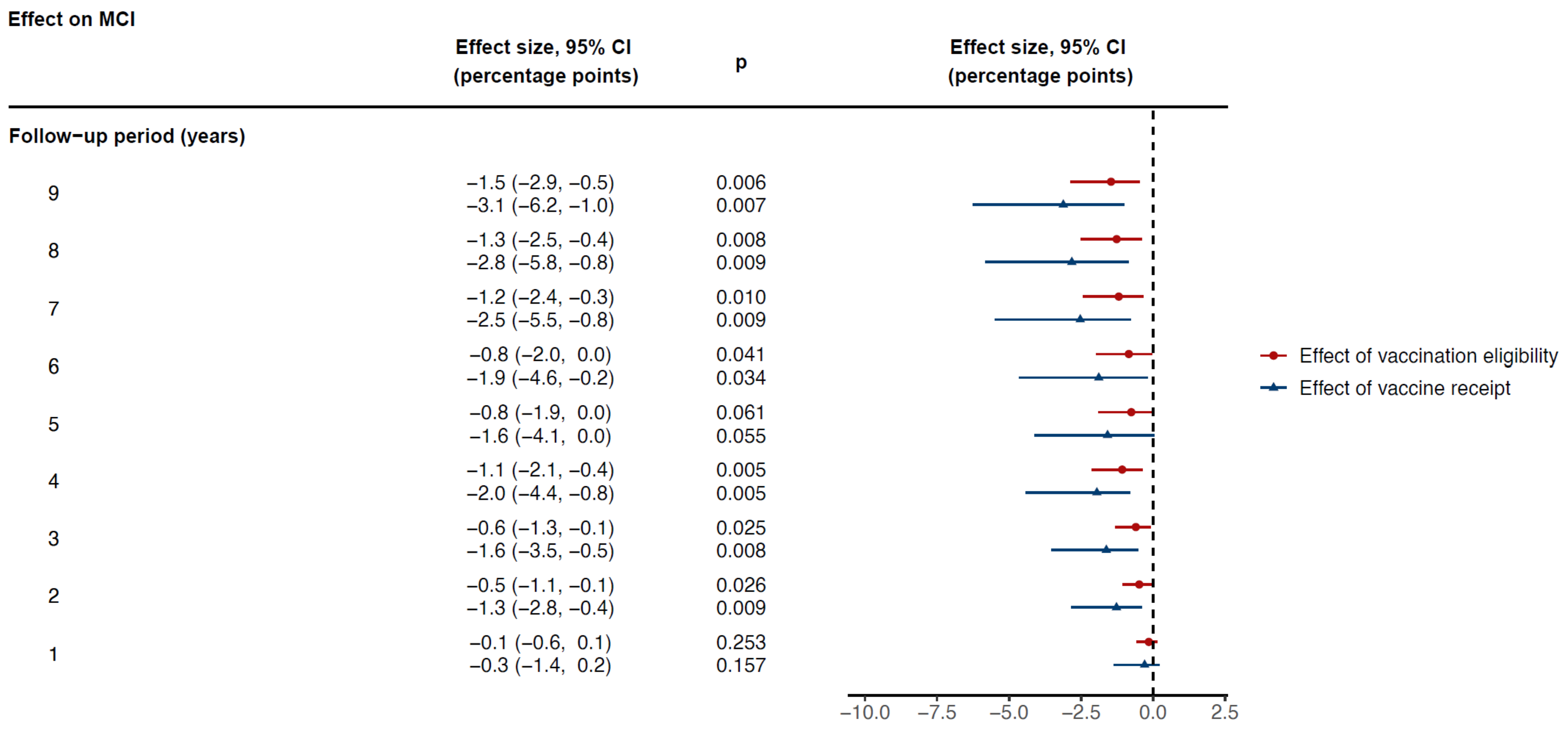


**Figure S6. The effect of HZ vaccination eligibility and receipt on new diagnoses of MCI across different lengths of follow-up.[1]**

[1] Dots or triangles show the point estimate and horizontal bars the 95% confidence interval.

Abbreviations: MCI = mild cognitive impairment; CI = confidence interval

**Effect on MCI**

| | Effect size, 95% CI (percentage points) | p |
|---|---|---|
| **Bandwidth choice (as x−times the MSE−optimal bandwidth)** | | |
| 2 | −0.9 (−1.7, −0.1) | 0.033 |
| | −1.9 (−4.0, −0.3) | 0.026 |
| 1.75 | −1.0 (−2.0, −0.2) | 0.017 |
| | −2.1 (−4.5, −0.5) | 0.014 |
| 1.5 | −1.1 (−2.3, −0.3) | 0.009 |
| | −2.4 (−4.9, −0.7) | 0.011 |
| 1.25 | −1.4 (−2.6, −0.5) | 0.005 |
| | −2.9 (−5.7, −1.0) | 0.005 |
| 1 | −1.5 (−2.9, −0.5) | 0.006 |
| | −3.1 (−6.2, −1.0) | 0.007 |
| 0.75 | −1.4 (−3.0, −0.2) | 0.027 |
| | −3.1 (−6.5, −0.2) | 0.036 |
| 0.5 | −1.7 (−3.4, 0.0) | 0.056 |
| | −3.7 (−8.3, −0.1) | 0.046 |
| **Grace period (months)** | | |
| 4 | −1.5 (−3.0, −0.5) | 0.005 |
| | −3.3 (−6.5, −1.2) | 0.004 |
| 2 | −1.5 (−2.9, −0.5) | 0.006 |
| | −3.1 (−6.3, −1.0) | 0.007 |
| 0 | −1.5 (−2.9, −0.5) | 0.006 |
| | −3.1 (−6.2, −1.0) | 0.007 |
| **Alternative outcome definition** | | |
| MCI not followed by dementia within 6 months | −1.3 (−2.6, −0.4) | 0.006 |
| | −2.8 (−5.6, −1.0) | 0.004 |
| MCI not followed by dementia within 3 months | −1.4 (−2.8, −0.5) | 0.006 |
| | −3.0 (−5.9, −1.1) | 0.005 |
| **Other checks** | | |
| Restricting to frequent GP visitors | −1.3 (−2.8, −0.3) | 0.017 |
| | −2.6 (−5.7, −0.4) | 0.022 |
| Quadratic fit | −1.7 (−3.3, −0.6) | 0.006 |
| | −3.5 (−6.7, −1.1) | 0.007 |
| Controlling for health service utilization | −1.4 (−2.8, −0.4) | 0.011 |
| | −2.9 (−6.0, −0.8) | 0.010 |
| Accounting for staggered roll−out | −1.5 (−2.8, −0.5) | 0.004 |
| | −3.3 (−6.8, −0.5) | 0.023 |

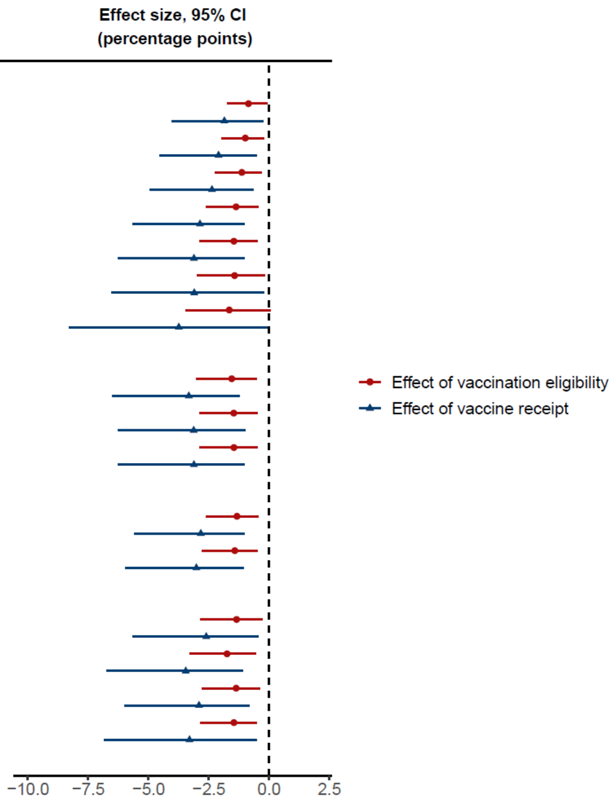


**Figure S7. Robustness checks for the effect of HZ vaccination eligibility and receipt on new diagnoses of MCI.**[1,2,3,4,5]

[1] Dots or triangles show the point estimate and horizontal bars the 95% confidence interval.

[2] With "grace periods" we refer to time periods since the index date after which follow-up time is considered to begin to allow for the time needed for a full immune response to develop after vaccine administration.

[3] Frequent GP visitors were defined as patients who had made at least one visit to their primary care provider during each of the five years preceding the start of the HZ vaccination program.

[4] The health service utilization indicators that were used when controlling for health service utilization were the number of primary care visits, outpatient visits, hospital admissions, and influenza vaccinations received during our nine-year follow-up period.

[5] As described in detail in the Methods section, when accounting for the staggered roll-out of the program, we adjusted the follow-up period to begin for each individual on the date on which they first became eligible for HZ vaccination (instead of starting the follow-up period for all individuals on September 1 2013). We added cohort fixed effects to these analyses to control for between-cohort differences in the date at which the follow-up window started. That is, we defined one cohort fixed effect for ineligible individuals and the first catch-up cohort, and included additional cohort fixed effects for each group of patients who became eligible at the same time.

Abbreviations: MCI = mild cognitive impairment; CI = confidence interval; MSE = mean squared error; GP = General Practitioner

**Effect on deaths due to dementia**

| Follow−up period (years) | Effect size, 95% CI (percentage points) | p |
|---|---|---|
| 9 | −8.5 (−18.5, −0.6) | 0.036 |
| | −29.5 (−62.9, −0.6) | 0.046 |
| 8 | −7.7 (−17.6, 0.2) | 0.056 |
| | −26.7 (−59.9, 2.1) | 0.068 |
| 7 | −8.2 (−18.5, −0.2) | 0.044 |
| | −28.4 (−63.3, 0.5) | 0.054 |
| 6 | −8.9 (−19.1, −1.0) | 0.029 |
| | −30.7 (−65.4, −2.1) | 0.036 |
| 5 | −6.6 (−15.3, 1.9) | 0.125 |
| | −22.8 (−52.1, 7.6) | 0.145 |
| 4 | −3.9 (−11.3, 3.8) | 0.334 |
| | −13.6 (−38.6, 13.9) | 0.356 |
| 3 | −3.4 (−10.7, 2.8) | 0.249 |
| | −12.0 (−37.0, 10.1) | 0.264 |
| 2 | −2.7 (−9.2, 3.7) | 0.407 |
| | −9.4 (−31.3, 13.3) | 0.430 |
| 1 | −2.6 (−7.1, 1.4) | 0.183 |
| | −8.9 (−24.2, 5.1) | 0.200 |

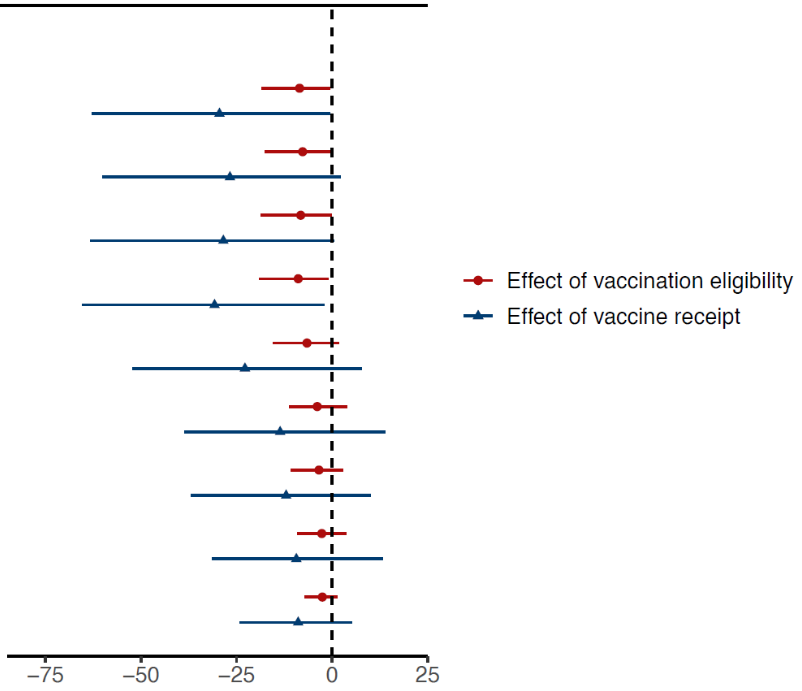


**Figure S8. The effect of HZ vaccination eligibility and receipt on deaths due to dementia across different lengths of follow-up.**[1]

[1] Dots or triangles show the point estimate and horizontal bars the 95% confidence interval.

Abbreviations: MCI = mild cognitive impairment; CI = confidence interval

**Effect on deaths due to dementia**

| | Effect size, 95% CI (percentage points) | p |
|---|---|---|
| **Bandwidth choice (as x−times the MSE−optimal bandwidth)** | | |
| 2 | −5.4 (−12.9, −0.4)<br>−20.6 (−50.1, −2.0) | 0.036<br>0.033 |
| 1.75 | −5.8 (−13.9, −0.5)<br>−22.0 (−53.1, −2.2) | 0.034<br>0.033 |
| 1.5 | −6.4 (−14.7, −0.2)<br>−23.5 (−54.2, −0.4) | 0.043<br>0.047 |
| 1.25 | −7.0 (−15.9, 0.0)<br>−24.7 (−55.5, 1.0) | 0.051<br>0.059 |
| 1 | −8.5 (−18.5, −0.6)<br>−29.5 (−62.9, −0.6) | 0.036<br>0.046 |
| 0.75 | −9.0 (−20.5, 0.2)<br>−32.2 (−73.1, 1.5) | 0.055<br>0.060 |
| 0.5 | −8.4 (−21.1, 4.7)<br>−29.3 (−73.9, 16.6) | 0.212<br>0.215 |
| **Grace period (months)** | | |
| 4 | −8.0 (−17.8, −0.1)<br>−27.9 (−60.7, 1.4) | 0.048<br>0.061 |
| 2 | −8.1 (−17.8, −0.2)<br>−28.0 (−60.7, 1.0) | 0.045<br>0.058 |
| 0 | −8.5 (−18.5, −0.6)<br>−29.5 (−62.9, −0.6) | 0.036<br>0.046 |
| **Other checks** | | |
| Quadratic fit | −8.9 (−19.6, 0.4)<br>−29.6 (−64.6, 2.7) | 0.061<br>0.072 |
| Adjusting for baseline IHD diagnosis | −14.3 (−28.0, −0.5) | 0.042 |
| Accounting for staggered roll−out | −8.1 (−18.0, −0.3)<br>−31.2 (−69.0, 0.0) | 0.043<br>0.050 |

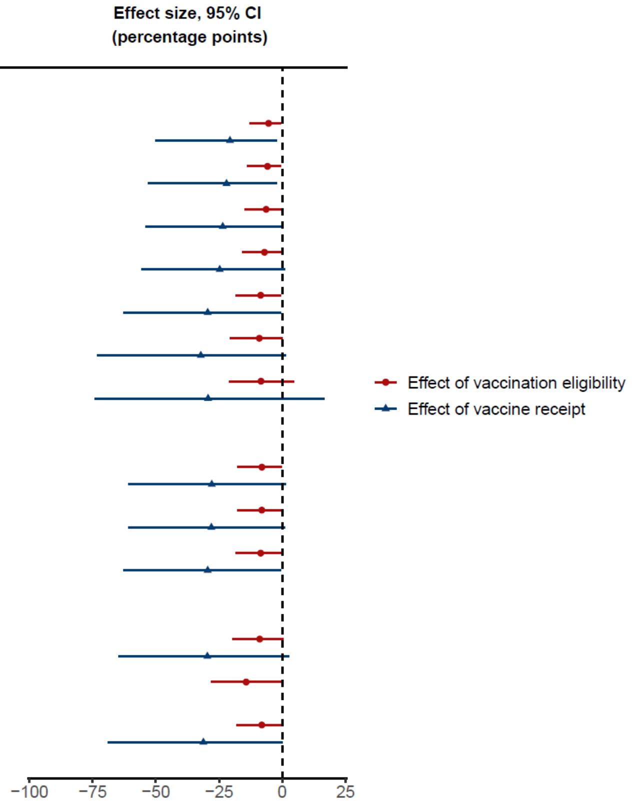


**Figure S9. Robustness checks for the effect of HZ vaccination eligibility and receipt on deaths due to dementia.**[1,2,3]

[1] Dots or triangles show the point estimate and horizontal bars the 95% confidence interval.

[2] With "grace periods" we refer to time periods since the index date after which follow-up time is considered to begin to allow for the time needed for a full immune response to develop after vaccine administration.

[3] As described in detail in the Methods, when accounting for the staggered roll-out of the program, we adjusted the follow-up period to begin for each individual on the date on which they first became eligible for HZ vaccination (instead of starting the follow-up period for all individuals on September 1 2013). We added cohort fixed effects to these analyses to control for between-cohort differences in the date at which the follow-up window started. That is, we defined one cohort fixed effect for ineligible individuals and the first catch-up cohort and included additional cohort fixed effects for each group of patients who became eligible at the same time.

Abbreviations: CI = confidence interval; MSE = mean squared error; IHD = ischemic heart disease

| | Effect size, 95% CI (percentage points) | p |
|---|---|---|
| **Eligibility cutoff with a 6−year follow−up** | | |
| 1936−09−02 | 0.3 (−0.5, 1.3) | 0.389 |
| | 0.2 (−7.5, 8.5) | 0.902 |
| 1935−09−02 | −0.1 (−0.8, 0.8) | 0.936 |
| | 0.8 (−7.8, 8.1) | 0.965 |
| 1934−09−02 | 0.2 (−0.6, 1.2) | 0.523 |
| | −0.7 (−7.4, 5.5) | 0.779 |
| 1933−09−02 | −0.9 (−2.0, 0.0) | 0.040 |
| | −8.9 (−19.1, −1.0) | 0.029 |
| 1932−09−02 | 0.3 (−0.4, 1.1) | 0.408 |
| | 3.8 (−4.9, 11.9) | 0.412 |
| 1931−09−02 | 0.0 (−1.0, 1.2) | 0.847 |
| | 3.4 (−4.9, 11.8) | 0.424 |
| 1930−09−02 | −0.5 (−2.1, 0.6) | 0.292 |
| | 2.2 (−5.4, 10.4) | 0.537 |

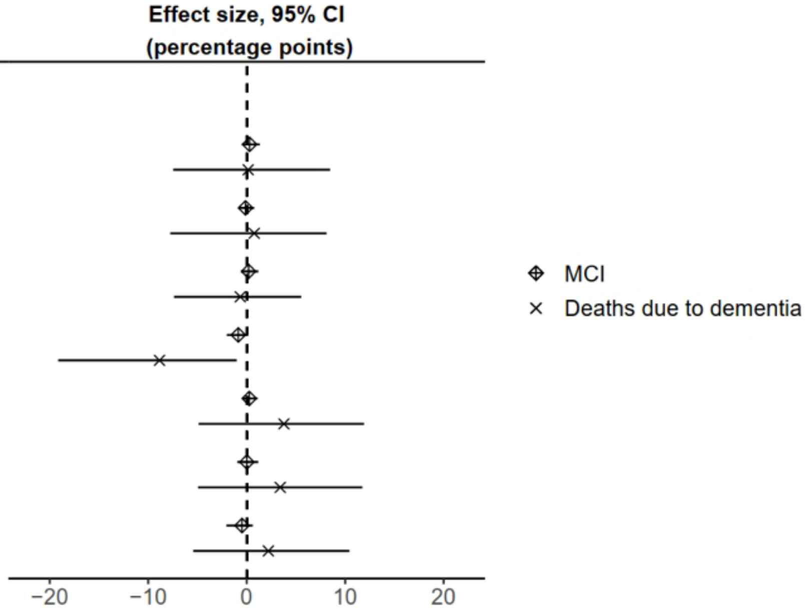


**Figure S10. The September 2 date-of-birth threshold only had a significant effect on our outcomes in the birth year (1933) that was used by the herpes zoster vaccination program as eligibility criterion.**[1,2,3,4]

[1] Diamonds or crosses show the point estimate and horizontal bars the 95% confidence interval.

[2] New diagnoses of MCI were analyzed among a study cohort of patients who did not have any record of cognitive impairment prior to the start of the follow-up period.

[3] Deaths due to dementia were analyzed among a study cohort of patients who had received a diagnosis of dementia prior to the start of the follow-up period.

[4] We used a six-year instead of a nine-year follow-up period in this analysis to allow for the same length of follow-up for all comparisons.

Abbreviations: CI = confidence interval; MCI = mild cognitive impairment

| | Effect size, 95% CI (percentage points) | p |
|---|---|---|
| **Incidence in the 9 years prior to Sept 1 2013** | | |
| MCI | 0.0 (–0.5, 0.6) | 0.953 |
| Deaths due to dementia | –5.2 (–17.7, 6.7) | 0.375 |

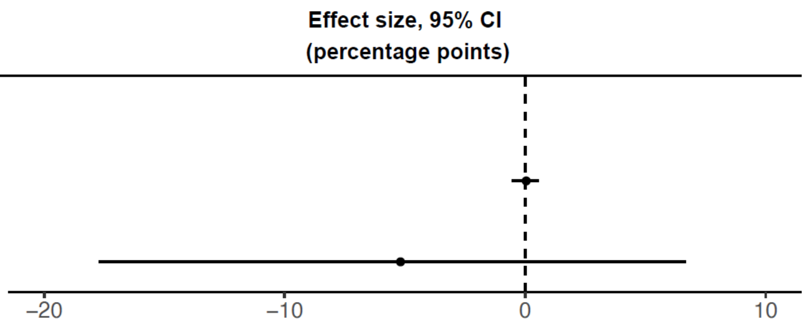


**Figure S11. The effect of the September 2 1933 eligibility threshold on diagnoses of MCI and deaths due to dementia occurring in the nine years prior to the start of the herpes zoster vaccination program.[1,2,3,4]**

[1] Dots show the point estimate and horizontal bars the 95% confidence interval.

[2] New diagnoses of MCI were analyzed among a study cohort of patients who did not have any record of cognitive impairment prior to September 1 2004.

[3] Deaths due to dementia were analyzed among a study cohort of patients who had received a diagnosis of dementia prior to September 1 2004.

[4] This figure shows the results from the identical analysis as implemented for our primary analysis (for which the results are shown in Fig. 2 in the main manuscript) except that we followed individuals from September 1 2004 to August 31 2013 instead of from September 1 2013 to August 31 2022. This analysis, thus, compared the exact same date-of-birth cohorts to each other and had the same length of follow-up as our primary analysis, but used the nine years prior to the start of the HZ vaccination program as follow-up period.

Abbreviations: CI = confidence interval; Sept = September; MCI = mild cognitive impairment

**Effect on MCI among women**

| | Effect size, 95% CI (percentage points) | p |
|---|---|---|
| **Bandwidth choice (as x−times the MSE−optimal bandwidth)** | | |
| 2 | −1.5 (−2.9, −0.5) | 0.004 |
| | −2.5 (−6.1, −0.9) | 0.008 |
| 1.75 | −1.7 (−3.3, −0.8) | 0.002 |
| | −3.1 (−7.1, −1.5) | 0.003 |
| 1.5 | −2.0 (−3.7, −1.0) | < 0.001 |
| | −3.6 (−8.0, −2.0) | 0.001 |
| 1.25 | −2.3 (−4.1, −1.1) | < 0.001 |
| | −4.3 (−8.8, −2.3) | < 0.001 |
| 1 | −2.5 (−4.5, −1.1) | 0.001 |
| | −5.1 (−9.8, −2.6) | < 0.001 |
| 0.75 | −2.6 (−4.8, −0.9) | 0.004 |
| | −5.6 (−10.7, −2.3) | 0.003 |
| 0.5 | −2.7 (−5.2, −0.3) | 0.026 |
| | −5.9 (−11.9, −1.0) | 0.021 |
| **Grace period (months)** | | |
| 4 | −2.5 (−4.6, −1.1) | 0.001 |
| | −5.3 (−10.1, −2.7) | < 0.001 |
| 2 | −2.4 (−4.4, −1.0) | 0.002 |
| | −4.9 (−9.6, −2.3) | 0.001 |
| 0 | −2.5 (−4.5, −1.1) | 0.001 |
| | −5.1 (−9.8, −2.6) | < 0.001 |
| **Alternative outcome definition** | | |
| MCI not followed by dementia within 6 months | −2.4 (−4.2, −1.1) | < 0.001 |
| | −4.8 (−9.1, −2.6) | < 0.001 |
| MCI not followed by dementia within 3 months | −2.5 (−4.5, −1.2) | < 0.001 |
| | −5.1 (−9.6, −2.8) | < 0.001 |
| **Other checks** | | |
| Restricting to frequent GP visitors | −2.4 (−4.5, −0.9) | 0.004 |
| | −4.3 (−9.0, −1.8) | 0.004 |
| Quadratic fit | −2.8 (−4.8, −1.2) | 0.001 |
| | −6.1 (−10.6, −2.5) | 0.001 |
| Controlling for health service utilization | −2.3 (−4.3, −1.0) | 0.002 |
| | −4.8 (−9.4, −2.3) | 0.001 |
| Accounting for staggered roll−out | −1.5 (−2.8, −0.5) | 0.004 |
| | −3.3 (−6.8, −0.5) | 0.023 |

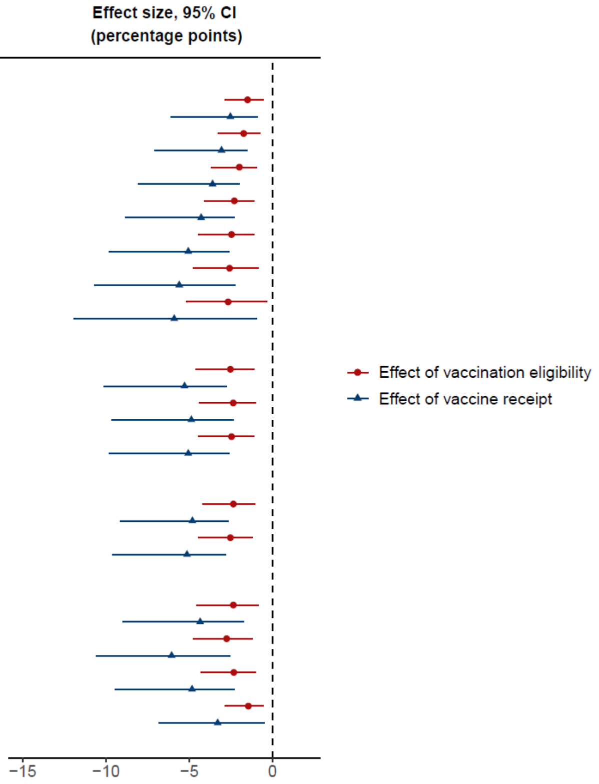


**Figure S12. Robustness checks for the effect of HZ vaccination eligibility and receipt on new diagnoses of MCI, among women only.**[1,2,3,4,5]

[1] Dots or triangles show the point estimate and horizontal bars the 95% confidence interval.

[2] With "grace periods" we refer to time periods since the index date after which follow-up time is considered to begin to allow for the time needed for a full immune response to develop after vaccine administration.

[3] Frequent GP visitors were defined as patients who had made at least one visit to their primary care provider during each of the five years preceding the start of the HZ vaccination program.

[4] The health service utilization indicators that were used when controlling for health service utilization were the number of primary care visits, outpatient visits, hospital admissions, and influenza vaccinations received during our nine-year follow-up period.

[5] As described in detail in the Methods section, when accounting for the staggered roll-out of the program, we adjusted the follow-up period to begin for each individual on the date on which they first became eligible for HZ vaccination (instead of starting the follow-up period for all individuals on September 1 2013). We added cohort fixed effects to these analyses to control for between-cohort differences in the date at which the follow-up window started. That is, we defined one cohort fixed effect for ineligible individuals and the first catch-up cohort, and included additional cohort fixed effects for each group of patients who became eligible at the same time.

Abbreviations: MCI = mild cognitive impairment; CI = confidence interval; MSE = mean squared error; GP = General Practitioner

Effect on deaths due to dementia among women

| | Effect size, 95% CI (percentage points) | p |
|---|---|---|
| **Bandwidth choice (as x−times the MSE−optimal bandwidth)** | | |
| 2 | −10.9 (−20.2, −4.3)<br>−45.4 (−85.0, −17.0) | 0.002<br>0.003 |
| 1.75 | −11.3 (−21.5, −4.5)<br>−46.4 (−88.7, −17.1) | 0.003<br>0.004 |
| 1.5 | −11.6 (−22.2, −3.8)<br>−46.5 (−89.0, −13.1) | 0.006<br>0.008 |
| 1.25 | −11.8 (−22.8, −2.5)<br>−45.5 (−86.7, −7.0) | 0.015<br>0.021 |
| 1 | −13.9 (−26.3, −3.4)<br>−52.3 (−97.9, −9.2) | 0.011<br>0.018 |
| 0.75 | −15.2 (−30.4, −3.8)<br>−57.9 (−117.0, −11.7) | 0.012<br>0.017 |
| 0.5 | −14.2 (−30.2, 2.8)<br>−52.3 (−111.9, 13.1) | 0.103<br>0.121 |
| **Grace period (months)** | | |
| 4 | −13.7 (−25.7, −3.4)<br>−52.0 (−96.7, −8.7) | 0.010<br>0.019 |
| 2 | −14.5 (−26.7, −4.2)<br>−55.0 (−100.3, −11.5) | 0.007<br>0.014 |
| 0 | −13.9 (−26.3, −3.4)<br>−52.3 (−97.9, −9.2) | 0.011<br>0.018 |
| **Other checks** | | |
| Quadratic fit | −14.0 (−27.5, −1.8)<br>−50.5 (−98.6, −3.4) | 0.026<br>0.036 |
| Adjusting for baseline IHD diagnosis | −16.2 (−28.5, −3.9) | 0.010 |
| Accounting for staggered roll−out | −12.9 (−25.0, −2.4)<br>−52.4 (−101.3, −6.4) | 0.017<br>0.026 |

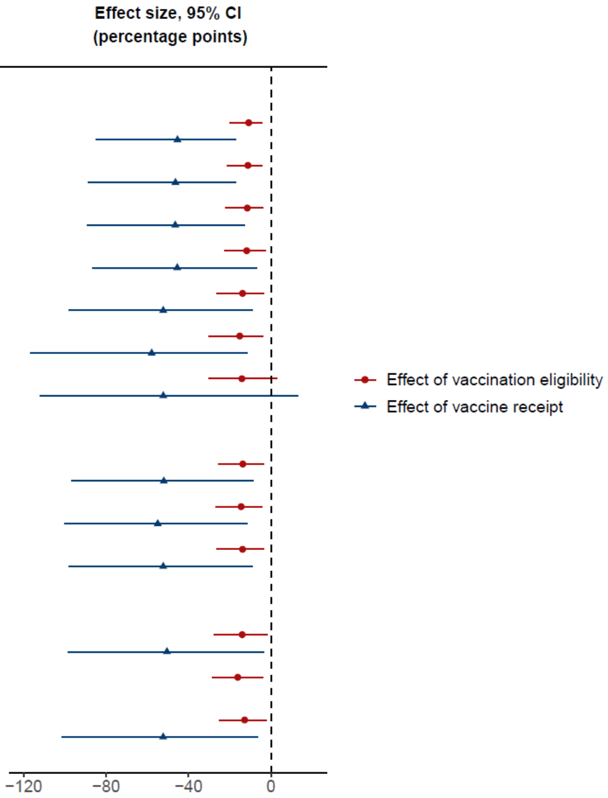


**Figure S13. Robustness checks for the effect of HZ vaccination eligibility and receipt on deaths due to dementia, among women only.**[1,2,3]

[1] Dots or triangles show the point estimate and horizontal bars the 95% confidence interval.

[2] With "grace periods" we refer to time periods since the index date after which follow-up time is considered to begin to allow for the time needed for a full immune response to develop after vaccine administration.

[3] As described in detail in the Methods section, when accounting for the staggered roll-out of the program, we adjusted the follow-up period to begin for each individual on the date on which they first became eligible for HZ vaccination (instead of starting the follow-up period for all individuals on September 1 2013). We added cohort fixed effects to these analyses to control for between-cohort differences in the date at which the follow-up window started. That is, we defined one cohort fixed effect for ineligible individuals and the first catch-up cohort, and included additional cohort fixed effects for each group of patients who became eligible at the same time.

Abbreviations: CI = confidence interval; MSE = mean squared error; IHD = ischemic heart disease

**Women**

| | Effect size, 95% CI (percentage points) | p |
|---|---|---|
| **Deaths due to dementia** | | |
| Effect of vaccine receipt | −52.3 (−97.9, −9.2)<br>−62.1 (−101.5, −22.7) | 0.018<br>0.002 |
| Effect of vaccination eligibility | −13.9 (−26.3, −3.4)<br>−14.9 (−23.9, −5.8) | 0.011<br>0.001 |
| **Non−dementia deaths** | | |
| Effect of vaccine receipt | 12.5 (−20.6, 45.4)<br>16.7 (−14.6, 47.9) | 0.462<br>0.296 |
| Effect of vaccination eligibility | 3.3 (−5.1, 12.0)<br>4.0 (−3.4, 11.3) | 0.434<br>0.288 |
| **All−cause mortality** | | |
| Effect of vaccine receipt | −34.7 (−63.0, −9.8)<br>−28.9 (−53.8, −3.9) | 0.007<br>0.023 |
| Effect of vaccination eligibility | −9.1 (−16.7, −3.1)<br>−6.9 (−12.8, −1.0) | 0.004<br>0.023 |

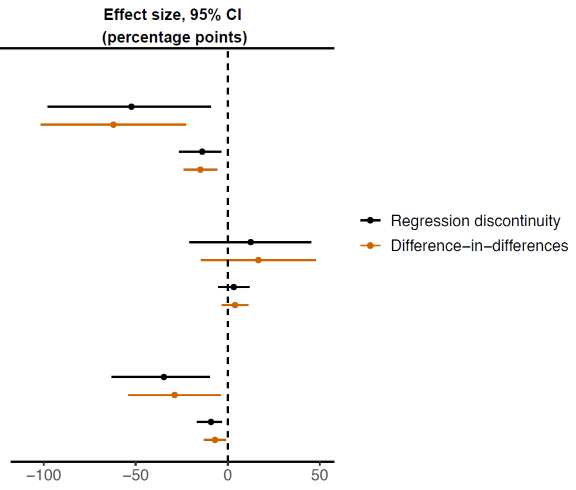


**Men**

| | Effect size, 95% CI (percentage points) | p |
|---|---|---|
| **Deaths due to dementia** | | |
| Effect of vaccine receipt | 4.3 (−40.1, 41.6)<br>1.4 (−38.9, 41.7) | 0.970<br>0.944 |
| Effect of vaccination eligibility | 1.3 (−12.4, 13.0)<br>0.4 (−11.0, 11.8) | 0.966<br>0.944 |
| **Non−dementia deaths** | | |
| Effect of vaccine receipt | −9.2 (−41.4, 24.0)<br>−13.4 (−49.2, 22.4) | 0.602<br>0.465 |
| Effect of vaccination eligibility | −2.7 (−12.0, 7.1)<br>−3.8 (−13.9, 6.3) | 0.614<br>0.462 |
| **All−cause mortality** | | |
| Effect of vaccine receipt | −4.1 (−29.7, 17.5)<br>−8.3 (−30.0, 13.3) | 0.613<br>0.450 |
| Effect of vaccination eligibility | −1.3 (−9.4, 5.5)<br>−2.4 (−8.6, 3.8) | 0.609<br>0.453 |

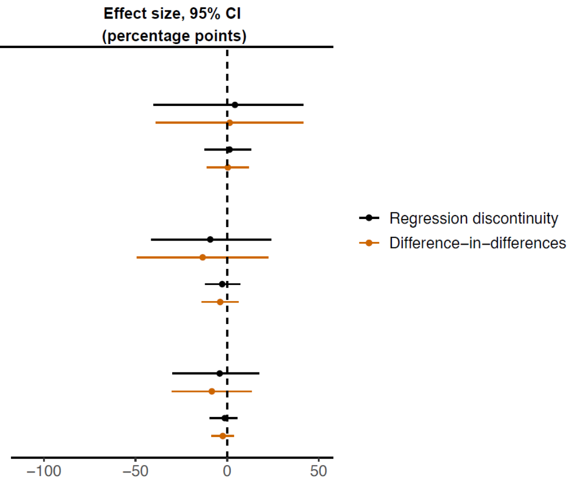


**Figure S14. The effect of HZ vaccination eligibility and receipt on deaths due to dementia, non-dementia deaths and all-cause mortality, separately among women and men.**[1,2,3]

[1] Dots show the point estimate and horizontal bars the 95% confidence interval.

[2] All outcomes were analyzed among a study cohort of patients who had received a diagnosis of dementia prior to the start date of the HZ vaccination program.

[3] Non-dementia deaths were defined as deaths for which dementia was neither the underlying nor a contributing cause of death in the death certificate.

Abbreviations: CI = confidence interval

**Effect on all−cause mortality**

| | Effect size, 95% CI (percentage points) | p |
|---|---|---|
| **Bandwidth choice (as x−times the MSE−optimal bandwidth)** | | |
| 2 | −4.3 (−8.6, −1.2)<br>−16.4 (−33.3, −4.9) | 0.010<br>0.008 |
| 1.75 | −4.4 (−9.1, −1.2)<br>−16.8 (−35.2, −5.1) | 0.010<br>0.009 |
| 1.5 | −4.6 (−9.7, −1.1)<br>−17.4 (−36.4, −4.4) | 0.014<br>0.013 |
| 1.25 | −5.3 (−10.6, −1.2)<br>−19.2 (−37.7, −4.0) | 0.014<br>0.015 |
| 1 | −6.5 (−12.5, −2.1)<br>−22.7 (−42.8, −6.5) | 0.006<br>0.008 |
| 0.75 | −8.1 (−15.5, −3.4)<br>−28.5 (−53.8, −11.1) | 0.002<br>0.003 |
| 0.5 | −9.1 (−17.5, −2.6)<br>−32.9 (−63.5, −9.6) | 0.008<br>0.008 |
| **Grace period (months)** | | |
| 4 | −8.4 (−15.7, −3.1)<br>−29.1 (−54.1, −9.2) | 0.003<br>0.006 |
| 2 | −7.2 (−13.6, −2.4)<br>−24.9 (−46.6, −7.3) | 0.005<br>0.007 |
| 0 | −6.5 (−12.5, −2.1)<br>−22.7 (−42.8, −6.5) | 0.006<br>0.008 |
| **Other checks** | | |
| Quadratic fit | −7.7 (−14.5, −2.5)<br>−25.9 (−47.5, −7.3) | 0.006<br>0.008 |
| Adjusting for baseline IHD diagnosis | −14.9 (−26.5, −3.3) | 0.012 |
| Accounting for staggered roll−out | −6.9 (−13.2, −2.4)<br>−26.6 (−50.8, −8.5) | 0.005<br>0.006 |

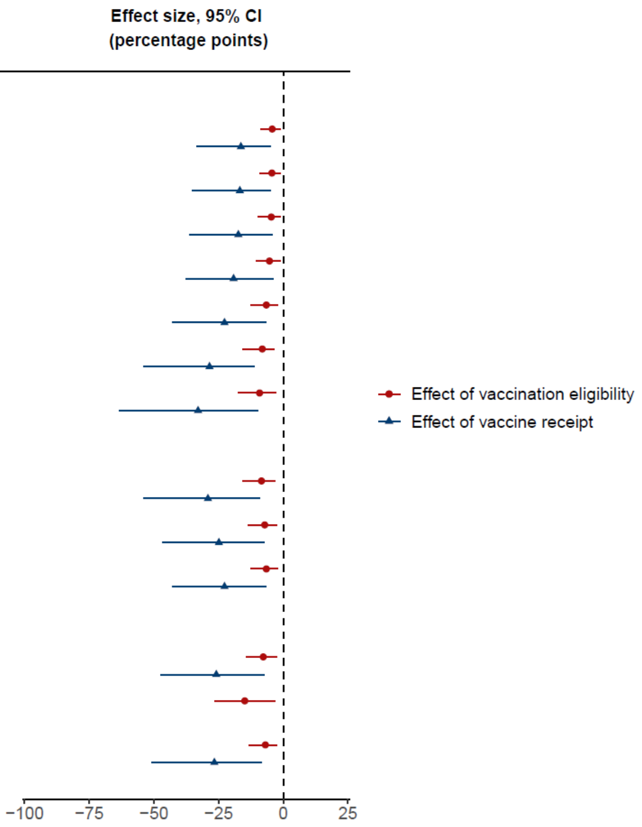


**Figure S15. Robustness checks for the effect of HZ vaccination eligibility and receipt on all-cause mortality among patients living with dementia at baseline.[1,2,3,4]**

[1] "Baseline" refers to the start date of the HZ vaccination program (i.e., September 1 2013).

[2] Dots or triangles show the point estimate and horizontal bars the 95% confidence interval.

[3] With "grace periods" we refer to time periods since the index date after which follow-up time is considered to begin to allow for the time needed for a full immune response to develop after vaccine administration.

[4] As described in detail in the Methods section, when accounting for the staggered roll-out of the program, we adjusted the follow-up period to begin for each individual on the date on which they first became eligible for HZ vaccination (instead of starting the follow-up period for all individuals on September 1 2013). We added cohort fixed effects to these analyses to control for between-cohort differences in the date at which the follow-up window started. That is, we defined one cohort fixed effect for ineligible individuals and the first catch-up cohort, and included additional cohort fixed effects for each group of patients who became eligible at the same time.

Abbreviations: CI = confidence interval; MSE = mean squared error; IHD = ischemic heart disease

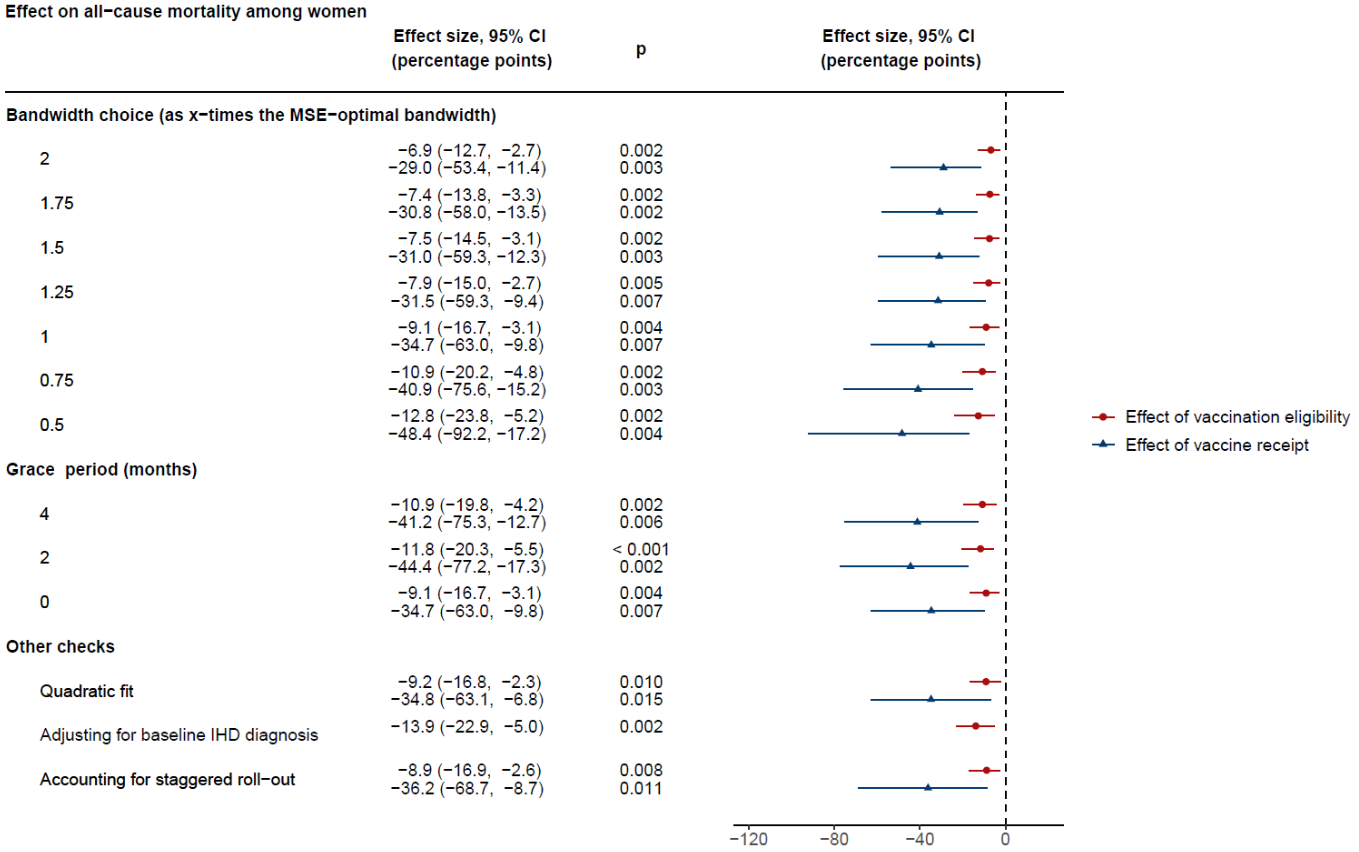


**Figure S16. Robustness checks for the effect of HZ vaccination eligibility and receipt on all-cause mortality among women living with dementia at baseline.**[1,2,3,4]

[1] “Baseline” refers to the start date of the HZ vaccination program (i.e., September 1 2013).

[2] Dots or triangles show the point estimate and horizontal bars the 95% confidence interval.

[3] With “grace periods” we refer to time periods since the index date after which follow-up time is considered to begin to allow for the time needed for a full immune response to develop after vaccine administration.

[4] As described in detail in the Methods section, when accounting for the staggered roll-out of the program, we adjusted the follow-up period to begin for each individual on the date on which they first became eligible for HZ vaccination (instead of starting the follow-up period for all individuals on September 1 2013). We added cohort fixed effects to these analyses to control for between-cohort differences in the date at which the follow-up window started. That is, we defined one cohort fixed effect for ineligible individuals and the first catch-up cohort, and included additional cohort fixed effects for each group of patients who became eligible at the same time.

Abbreviations: CI = confidence interval; MSE = mean squared error; IHD = ischemic heart disease

| | Mild Cognitive Impairment<br>(1) | Deaths due to dementia<br>(2) |
|---|---|---|
| Difference in CACE by gender | 4.9 | 55.9 |
| 95% CI | (-3.3, 6.5) | (35.4, 76.3) |
| p | 0.029 | 0.039 |
| Bandwidth (in weeks) | 95.14 | 97.5 |
| Observations | 282,557 | 14,350 |

**Table S1. Difference (in percentage points) between women and men in the effect of receipt of HZ vaccination on new diagnoses of mild cognitive impairment and deaths due to dementia.**[1]

[1] The CACE (complier average causal effect) refers to the estimated effect of actually receiving HZ vaccination as opposed to merely being eligible for vaccination. The reference group was women when calculating the difference in CACE. Bandwidth refers to the mean squared error-optimal bandwidth. Observations refer to the number of observations within the mean squared error-optimal bandwidth.

Abbreviations: CACE = complier average causal effect; CI = confidence interval; p = p-value

15. Office for National Statistics, Impact of registration delays on mortality statistics in England and Wales - Office for National Statistics. https://www.ons.gov.uk/peoplepopulationandcommunity/birthsdeathsandmarriages/deaths/articles/impactofregistrationdelaysonmortalitystatisticsinenglandandwales/latest.

16. Office for National Statistics (ONS), User guide to mortality statistics (2022). https://www.ons.gov.uk/peoplepopulationandcommunity/birthsdeathsandmarriages/deaths/methodologies/userguidetomortalitystatisticsjuly2017.

17. G. W. Imbens, T. Lemieux, Regression discontinuity designs: A guide to practice. *Journal of Econometrics* **142**, 615–635 (2008).

18. Global Burden of Disease Project 2019, GBD Compare (2020). http://vizhub.healthdata.org/gbd-compare.
